# Spatial machine learning and longitudinal analysis of skilled antenatal care access and fertility-related inequities in Ghana (1988–2022)

**DOI:** 10.64898/2026.07.16.26358217

**Authors:** Valentine Golden Ghanem

## Abstract

This study focuses on the relationship between access to Advanced Neonatal Care (ANC) and fertility across the regions in Ghana between 1988 and 2022. It builds on previous studies focused on inequity in maternal health across subnational levels and incorporates spatial analytics, machine learning, and a welfare-adjusted fertility care metric.

Nine waves of the Ghana Demographic and Health Survey (DHS) were analyzed, with 94 region-by-year units across 8 to 16 regions in each survey wave in the 16 Ghana administrative regions. Skilled ANC along with the Total Fertility Rate (TFR) and demographic control variables were extracted for the analysis. The methodologies employed include decomposition of the Gini coefficient of inequality, bivariate z-score risk stratification, Random Forest (RF), and Decision Tree (DT) regression, partial dependence, Local Indicators of Spatial Association (LISA), global Moran’s I with permutation inference and a novel Care Efficiency Index (CEI = ANC% / TFR). Care for the outcomes employed region aggregations along with district boundary geometries for the display of the choropleth maps.

National skilled ANC coverage increased from 83.1% (1988) to 97.7% (2022), with inter-regional Gini declining 87.9% (0.070 to 0.008). The North-South gap narrowed from 32.4 to 0.9 percentage points. Northern region showed the greatest absolute gain (+43.0pp). Machine learning identified an exploratory RF partial-dependence inflection near TFR=5.90, above which predicted ANC coverage declined in the historical data. Survey year was the dominant RF predictor (43.7%), followed by TFR (38.8%). TFR spatial clustering intensified by 2022 (Moran’s I=0.606, p=0.001). Greater Accra led the Care Efficiency Index (CEI=31.9); Northern Belt regions lagged (CEI=14.5-16.5). Risk stratification classified 23 observations as Critical (Low ANC/High TFR), predominantly from Northern Belt regions in earlier survey waves.

ANC coverage converged substantially, yet fertility-related spatial inequities persisted, especially in the Northern Belt. The Care Efficiency Index and exploratory TFR inflection provide hypothesis-generating tools for targeting health-system investment. They should not be interpreted as causal thresholds.

## 1. INTRODUCTION

Skilled antenatal care (ANC) is among the most cost-effective preventive interventions for reducing maternal and neonatal mortality in low- and middle-income countries (LMICs) [1]. The World Health Organization recommends a minimum of eight ANC contacts throughout pregnancy, with attendance by a skilled provider at every visit [1]. Despite substantial global progress, sub-Saharan Africa continues to bear a disproportionate share of the global maternal mortality burden—accounting for approximately 66% of worldwide maternal deaths—with coverage deficits concentrated in the highest-fertility, most geographically marginalised communities [2,16].

Ghana is an instructive setting for studying ANC scale-up. Since the late 1980s, the country has moved from under 60% skilled ANC coverage in the lowest-performing regions to near-universal coverage by 2022. This transition occurred alongside National Health Insurance Scheme (NHIS) expansion, the 2003 fee-exemption policy for facility-based delivery services, and community health worker programme scale-up [14,15]. Nevertheless, subnational variation in ANC access and total fertility rate (TFR) persists across Ghana’s DHS regional analytic units [3,19]. These patterns reflect structural disparities in health infrastructure, geographic access, insurance enrolment, female educational attainment, and community-level sociocultural norms [4,5].

This analysis builds on the broader Ghana ANC spatial-equity programme by shifting from district-level insurance inequity to the longitudinal relationship between skilled ANC coverage and fertility. Nine waves of Ghana Demographic and Health Survey (DHS) subnational data (1988-2022) were used to characterise the evolving spatial and temporal dynamics of ANC access in relation to fertility across all 16 Ghana administrative regions.

The relationship between TFR and ANC coverage is theoretically and empirically complex. In very high-fertility settings, compounding cultural, logistical, and economic barriers—including partner opposition, distance to facilities, and transportation costs—may suppress ANC utilisation independently of supply-side determinants [4,7]. Disentangling this relationship across time and space requires methods that transcend traditional regression, encompassing spatial autocorrelation analysis, machine learning feature attribution, and novel composite performance metrics.

The Care Efficiency Index (CEI = ANC coverage / TFR) was introduced as an exploratory region-specific composite. The index captures how high-fertility settings convert need for maternal health contact into skilled antenatal contact and complements established frameworks for monitoring reproductive health system performance [14,18]. Region-year observations were also classified into bivariate risk zones (Critical, Emerging, Workhorse, Resilient) and use partial dependence analysis to identify a non-linear TFR inflection associated with lower predicted ANC coverage [10,11].

This study had four objectives: to characterise the spatiotemporal epidemiology of skilled ANC access and fertility-related inequities in Ghana from 1988 to 2022; to quantify inter-regional inequality trajectories using the Gini coefficient [12]; to identify predictors of ANC coverage using machine learning and partial dependence analysis [10,11]; and to develop spatial typologies that can guide targeted maternal health investment.

## 2. METHODS

### 2.1 Study Design and Setting

This was a longitudinal ecological study at the subnational (regional) level in Ghana, covering nine DHS survey waves: 1988, 1993, 1998, 2003, 2008, 2014, 2016, 2019, and 2022. The analytical unit was the region-year observation, yielding 94 region-year observations across all 16 Ghana administrative regions, including Bono East and Western North (created via the 2018 administrative subdivision), for which independent 2022 DHS outcome rows are available. Reporting follows the Strengthening the Reporting of Observational Studies in Epidemiology (STROBE) guidelines for ecological studies.

### 2.2 Data Sources

Data were obtained from the Ghana Demographic and Health Surveys (DHS) subnational database (ICF International, Rockville, MD) [3,13]. Preferred Estimates (IsPreferred = TRUE) were extracted for two indicator domains: (1) antenatal care access indicators (including "Antenatal care from a skilled provider" and "No antenatal care"), and (2) fertility indicators (including "Total fertility rate 15–49" and age-specific rates). District-level coordinates (latitude/longitude centroids for 261 MMDAs) were extracted from the 2021 Ghana District Assembly administrative dataset.

### 2.3 Variable Definitions

The primary outcome was Skilled ANC Coverage (%), defined as the percentage of live births where antenatal care was received from a skilled provider (doctor, nurse, or midwife). The primary exposure was Total Fertility Rate (TFR), defined as the estimated number of live births per woman aged 15–49 years over her lifetime. Covariates included survey year, geographic zone (Northern/Middle/Southern Belt), adolescent fertility rate (15–19 years), and district-level poverty, illiteracy, and uninsurance rates from the Master Sheet. The Care Efficiency Index (CEI) was computed as CEI = Skilled ANC (%) / TFR for each region-year observation, providing a measure of service utilisation efficiency relative to fertility burden.

### 2.4 Inequality Analysis

The Gini coefficient was used to measure inequality between ANC (Antenatal Care) data across regions [12]. The coefficient is measured between 0 (complete equality) and 1 (complete inequality). For the analysis of North-South inequality, three defined regions, or “belts,” were used: Northern Belt (Northern, Upper East, Upper West, Savannah, North East), Middle Belt (Ashanti, Ahafo, Bono), and Southern Belt (Greater Accra, Central, Eastern, Western, Volta, Oti).

### 2.5 Spatial Autocorrelation Analysis

Global Moran’s I [8] was used to measure global spatial autocorrelation with k-nearest neighbor (KNN) spatial weights with k=4 based on the centroids of the regions. A permutation test with 999 iterations was used to facilitate the placement of the spatial weights. Local Indicators of Spatial Association (LISA) [9] was used to measure boundary adjacency with Rook contiguity (shared-boundary adjacency) weights. A significance level of p<0.10 was used based on 999 permutations. The classification of the LISA quadrants was based on the High-High (HH), Low-Low (LL), High-Low (HL), and Low-High (LH) standards. The Getis-Ord Gi* [27] statistic was considered as a local hotspot statistic, but was not used, given that LISA’s quadrant classification is more aligned with the established bivariate risk-stratification framework in Section 2.7. The choropleth maps used dissolved district-boundary geometry for display, while the DHS (Demographic and Health Surveys) estimates remained regional aggregates [22].

### 2.6 Machine Learning Modelling

Two supervised learning models were applied to predict skilled ANC coverage: a Decision Tree Regressor (DT) and a Random Forest Regressor (RF; 200 estimators, max_depth=6) [10]. Features included TFR, survey year, geographic zone, and adolescent fertility rate. The complete-case machine-learning subset was split 80:20 (train:test) with stratification by survey year. Model performance was evaluated using R2, root mean square error (RMSE), and mean absolute error (MAE). Five-fold cross-validation was applied to the RF model to assess generalisation [10]. Partial dependence analysis was conducted using the method of Friedman [11] to identify the TFR value at which predicted ANC coverage exhibited maximum gradient decline. All models were implemented using scikit-learn [23].

### 2.7 Risk Stratification

A bivariate z-score risk stratification was applied to all region-year observations. ANC z-scores and TFR z-scores were computed relative to the grand mean and standard deviation across all observations. Four zones were defined: Critical (ANC z<0 and TFR z>0; low ANC/high TFR), Emerging (ANC z<0 and TFR z<=0), Workhorse (ANC z>=0 and TFR z>0), and Resilient (ANC z>=0 and TFR z>=0).

### 2.8 Statistical and Computational Environment

All analyses were conducted in Python 3.12 using pandas [24], NumPy, scikit-learn [23], SciPy, GeoPandas, and libpysal/esda for spatial statistics [8,9]. Figures for submission were exported separately at 300 DPI or higher; analysis figures were generated with Matplotlib [25]. Early exploratory dashboards used Plotly [26] before migrating to the bespoke, dependency-free HI-EI interactive dashboard and poster, built as self-contained HTML artefacts with inlined visual assets. The aggregate analytic dataset, code, dashboard, and poster are available in the project repository.

Post hoc sensitivity analyses were conducted to test whether the main conclusions depended on modelling choices. Risk-zone labels were recomputed using same-year z-scores rather than the grand historical mean and standard deviation. ANC-TFR associations were re-estimated using heteroskedasticity-robust ordinary least squares, region-clustered and survey-year-clustered standard errors, region and survey-wave fixed effects, and generalized estimating equations with exchangeable region-level correlation. The pooled regional CEI gap was stress-tested using a descriptive bootstrap over regional mean CEI values. These analyses are reported as Supplementary File S1.

### 2.9 Ethical Considerations

This study used publicly available, de-identified, aggregate subnational data from the DHS programme [13]. No individual-level data were used. Institutional ethics review was not required per applicable guidelines for secondary analysis of publicly available anonymised data.

## 3. RESULTS

### 3.1 Phase I: Temporal and Regional Trends in ANC Coverage

National skilled ANC coverage increased from 83.1% (SD=13.0) in 1988 to 97.7% (SD=1.6) in 2022, representing a 17.5 percentage-point absolute gain over 34 years. The rate of improvement was most rapid between 1988 and 2003 (+8.8pp) and then plateaued as coverage approached saturation—a pattern consistent with ceiling effects observed as countries approach universal coverage targets. Complete regional-level ANC and fertility data are presented in Table 1; national summary statistics across all survey waves are shown in Table 2. Fig 1 provides a spatio-temporal heatmap of ANC coverage across all 16 Ghana administrative regions and nine survey waves.

**Fig 1.**
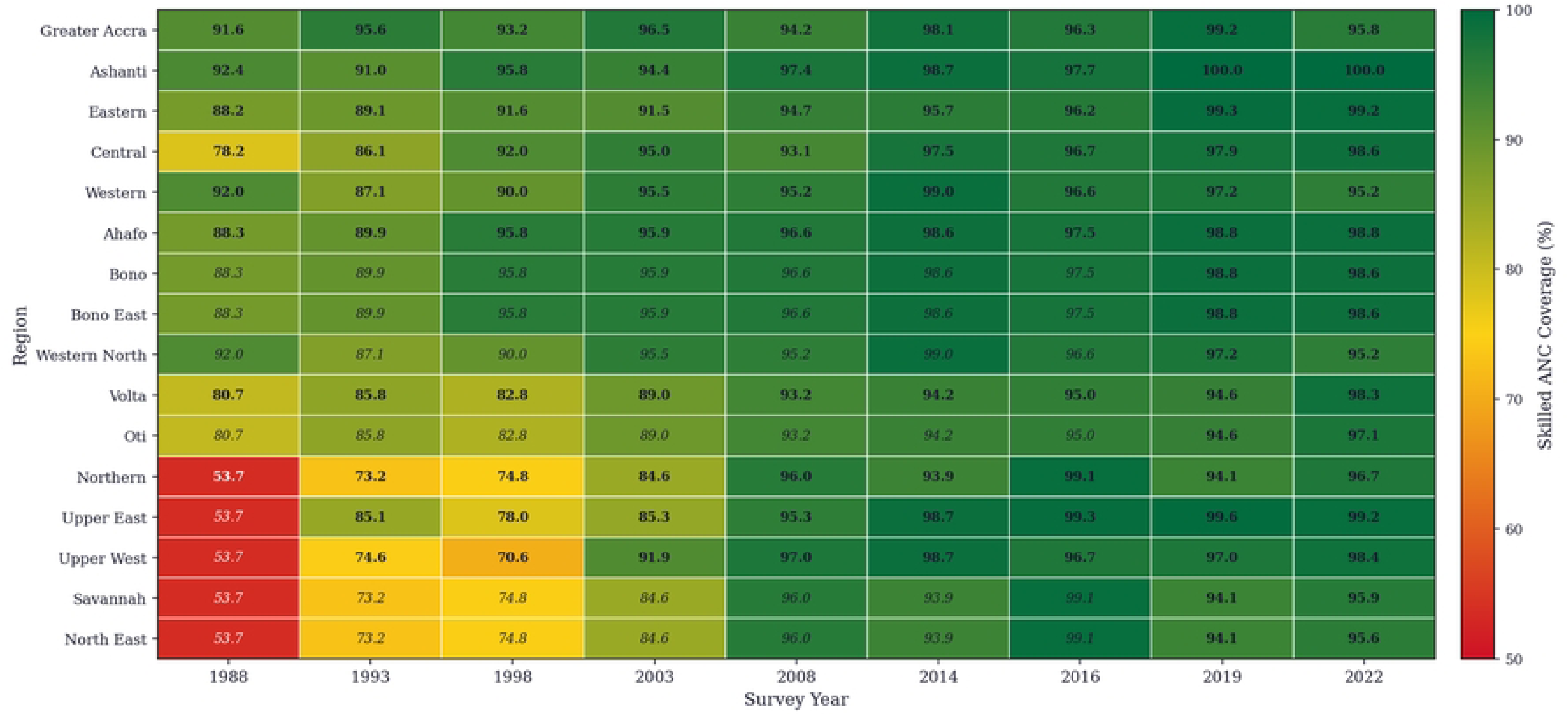
Spatio-temporal heatmap of skilled ANC coverage (%) by region and survey year, Ghana 1988–2022. Colour scale: red = low coverage, yellow = moderate, green = high coverage. Bold values = direct survey data; italic = propagated estimates from historical parent region following administrative subdivision. Source: Ghana DHS 1988–2022 [3,13].

**Table 1.** Regional Skilled ANC Coverage and Fertility Change, Ghana 1988–2022.

| Region | First wave | Last wave | ANC first (%) | ANC last (%) | Abs. change (pp) | % change | TFR first | TFR last |
| --- | --- | --- | --- | --- | --- | --- | --- | --- |
| Ahafo | 1988 | 2022 | 88.3 | 98.8 | +10.5 | +11.9% | 7.6 | 4.2 |
| Ashanti | 1988 | 2022 | 92.4 | 100.0 | +7.6 | +8.2% | 6.3 | 3.5 |
| Bono | 2022 | 2022 | 98.6 | 98.6 | +0.0 | +0.0% | 3.7 | 3.7 |
| Bono East | 2022 | 2022 | 99.4 | 99.4 | +0.0 | +0.0% | 4.7 | 4.7 |
| Central | 1988 | 2022 | 78.2 | 98.6 | +20.4 | +26.1% | 6.6 | 3.6 |
| Eastern | 1988 | 2022 | 88.2 | 99.2 | +11.0 | +12.5% | 6.0 | 3.5 |
| Greater Accra | 1988 | 2022 | 91.6 | 95.8 | +4.2 | +4.6% | 4.7 | 2.9 |
| North East | 2022 | 2022 | 95.6 | 95.6 | +0.0 | +0.0% | 6.6 | 6.6 |
| Northern | 1988 | 2022 | 53.7 | 96.7 | +43.0 | +80.1% | 6.9 | 5.4 |
| Oti | 2022 | 2022 | 97.1 | 97.1 | +0.0 | +0.0% | 5.2 | 5.2 |
| Savannah | 2022 | 2022 | 95.9 | 95.9 | +0.0 | +0.0% | 5.8 | 5.8 |
| Upper East | 1993 | 2022 | 85.1 | 99.2 | +14.1 | +16.6% | 5.7 | 4.6 |
| Upper West | 1993 | 2022 | 74.6 | 98.4 | +23.8 | +31.9% | 5.2 | 4.5 |
| Volta | 1988 | 2022 | 80.7 | 98.3 | +17.6 | +21.8% | 7.2 | 3.9 |
| Western | 1988 | 2022 | 92.0 | 95.2 | +3.2 | +3.5% | 6.4 | 3.7 |
| Western North | 2022 | 2022 | 98.6 | 98.6 | +0.0 | +0.0% | 3.8 | 3.8 |

pp = percentage points. ANC = antenatal care. TFR = total fertility rate.

**Table 2.** Spatial-Temporal Evolution of ANC and Fertility Indicators, Ghana 1988–2022.

| Year | N Regions | ANC<br>Mean (%) | ANC SD | TFR<br>Mean | TFR SD | TFR–ANC<br>Correlation | Gini (ANC) |
| --- | --- | --- | --- | --- | --- | --- | --- |
| 1988 | 8 | 83.1 | 13.0 | 6.46 | 0.88 | –0.360 | 0.070 |
| 1993 | 10 | 85.8 | 7.0 | 5.16 | 0.71 | –0.659 | 0.041 |
| 1998 | 10 | 86.5 | 9.2 | 4.63 | 0.86 | –0.459 | 0.055 |
| 2003 | 10 | 92.0 | 4.4 | 4.62 | 0.83 | –0.537 | 0.025 |
| 2008 | 10 | 95.3 | 1.5 | 4.20 | 0.95 | +0.113 | 0.009 |
| 2014 | 10 | 97.3 | 2.0 | 4.46 | 0.86 | –0.328 | 0.010 |
| 2016 | 10 | 97.1 | 1.3 | 4.20 | 0.68 | +0.553 | 0.007 |
| 2019 | 10 | 97.8 | 2.1 | 3.98 | 0.70 | –0.594 | 0.011 |
| 2022 | 16 | 97.8 | 1.5 | 4.35 | 0.99 | –0.387 | 0.008 |

SD = standard deviation. Gini = inter-regional Gini coefficient for ANC coverage.

At the regional level, Northern region recorded the largest absolute gain (+43.0pp; 53.7% to 96.7%), followed by Upper West (+23.8pp; 74.6% to 98.4%) and Central (+20.4pp; 78.2% to 98.6%). Greater Accra, already a high performer in 1988 (91.6%), showed the smallest long-run absolute change among regions observed from 1988 (+4.2pp), reflecting ceiling effects (Table 1). Regional trajectories by geographic belt are shown in Fig 2, and the TFR-ANC cross-sectional relationship across all region-year observations is shown in Fig 3.

**Fig 2.**
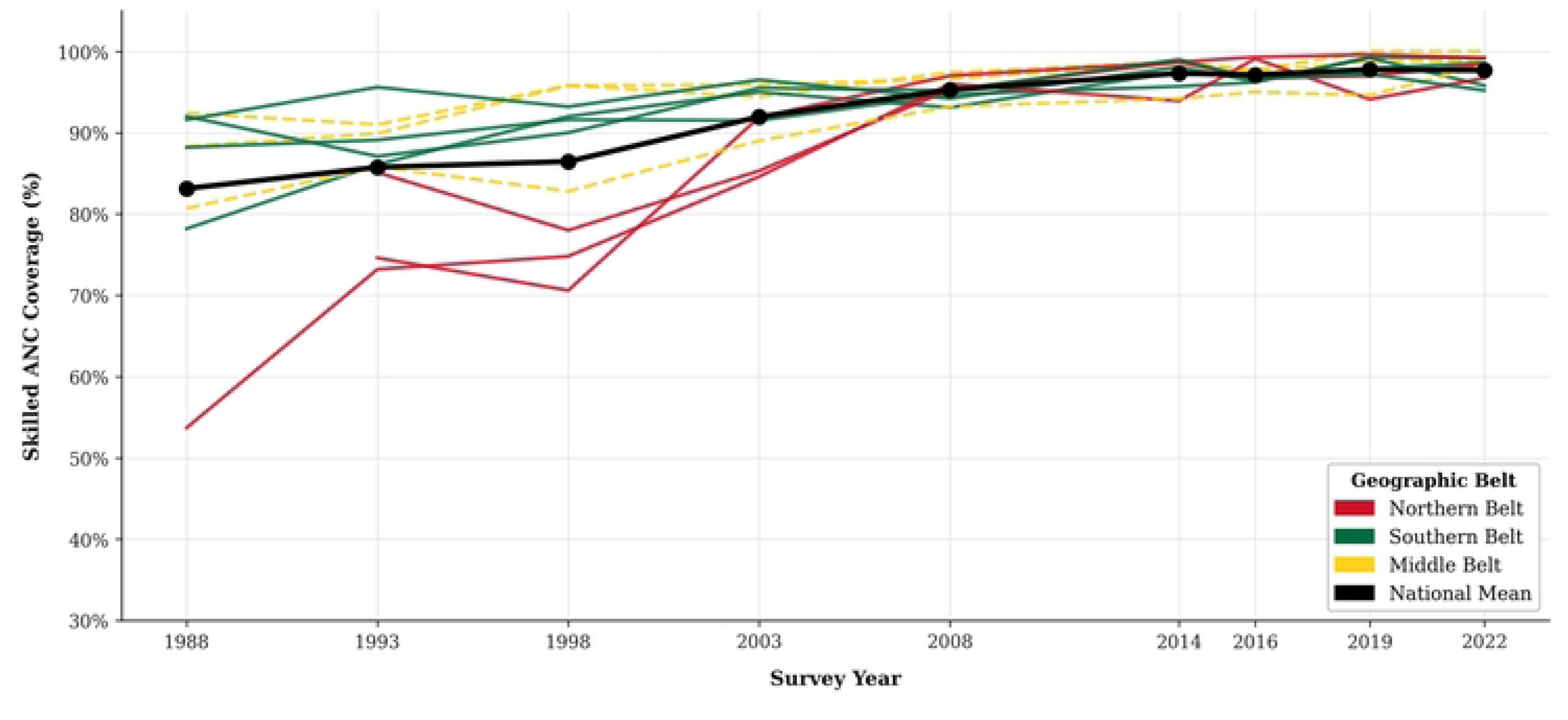
Regional skilled ANC coverage trajectories by geographic belt, Ghana 1988–2022. Northern Belt (red); Middle Belt (gold); Southern Belt (green). Each line represents one region. Legend identifies individual regions within each belt. Source: Ghana DHS 1988-2022 (Subnational, Preferred Estimates). Belt. Northern = Upper East/West, North East, Northern, Savannah; Southern = Greater Accra, Central, Western, Eastern, Middle = remaining regions.

**Fig 3.**
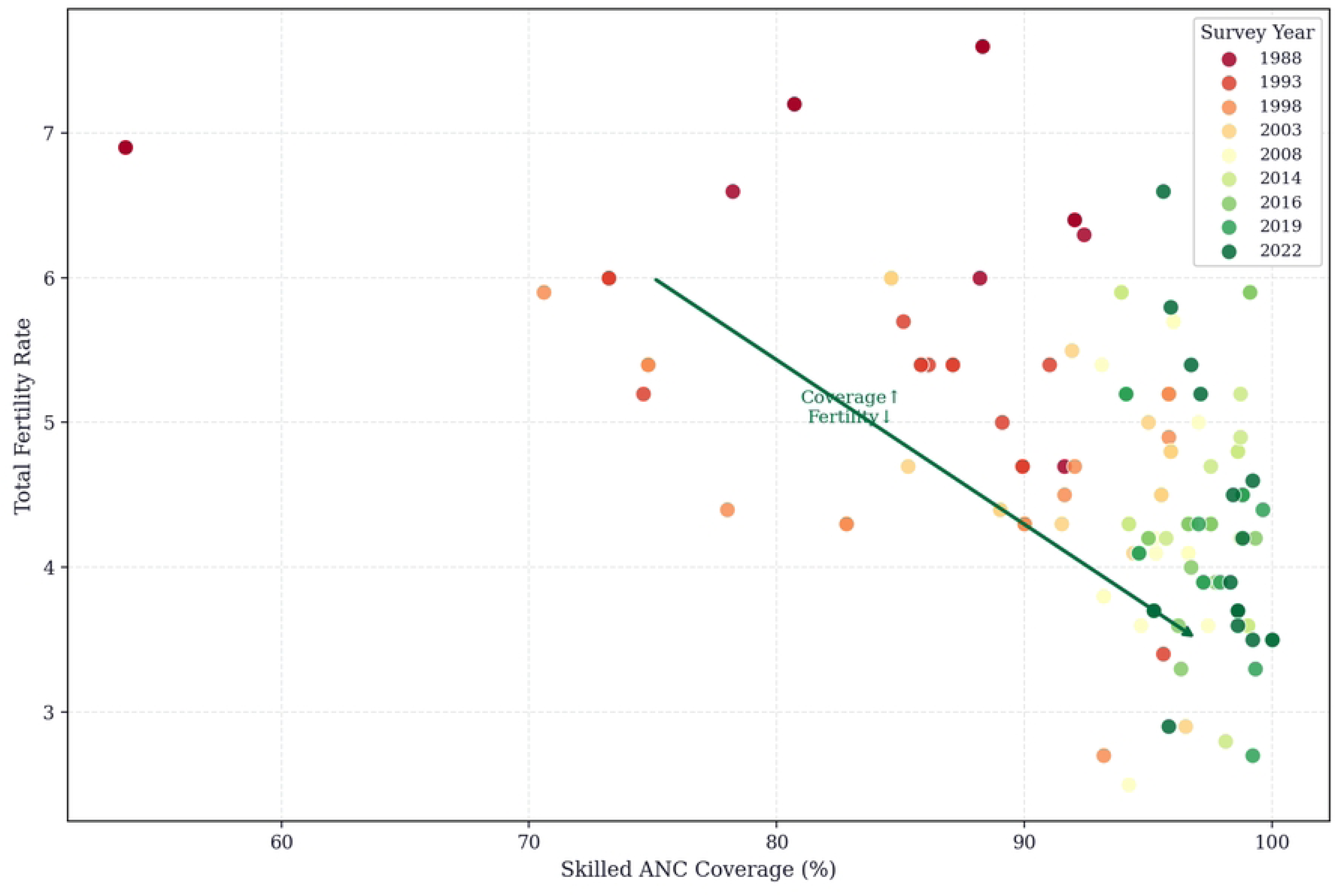
Scatter plot of regional TFR versus skilled ANC coverage, Ghana 1988–2022. Each observation is listed by region for the selected year with the observation year denoted by the color of the marker. An inverse relationship between total fertility rate and skilled ANC coverage exists for the majority of the years.

### 3.2 Phase II: Inequality and North–South ANC Disparities

Inter-regional inequality in ANC coverage declined 87.9%, from 0.070 in 1988 to 0.008 in 2022 (Fig 4). This pattern indicates convergence of Northern Belt regions toward Southern Belt levels. The North-South ANC gap narrowed from 32.4 percentage points in 1988 to under 1 percentage point by 2022. This magnitude is consistent with coverage improvements reported after fee-exemption policy reforms in comparable LMIC settings [14,15]. In contrast, the TFR Gini declined by only 27% over the same period (Fig 5), suggesting that fertility inequality has been more resistant to policy intervention than service coverage inequality.

**Fig 4.**
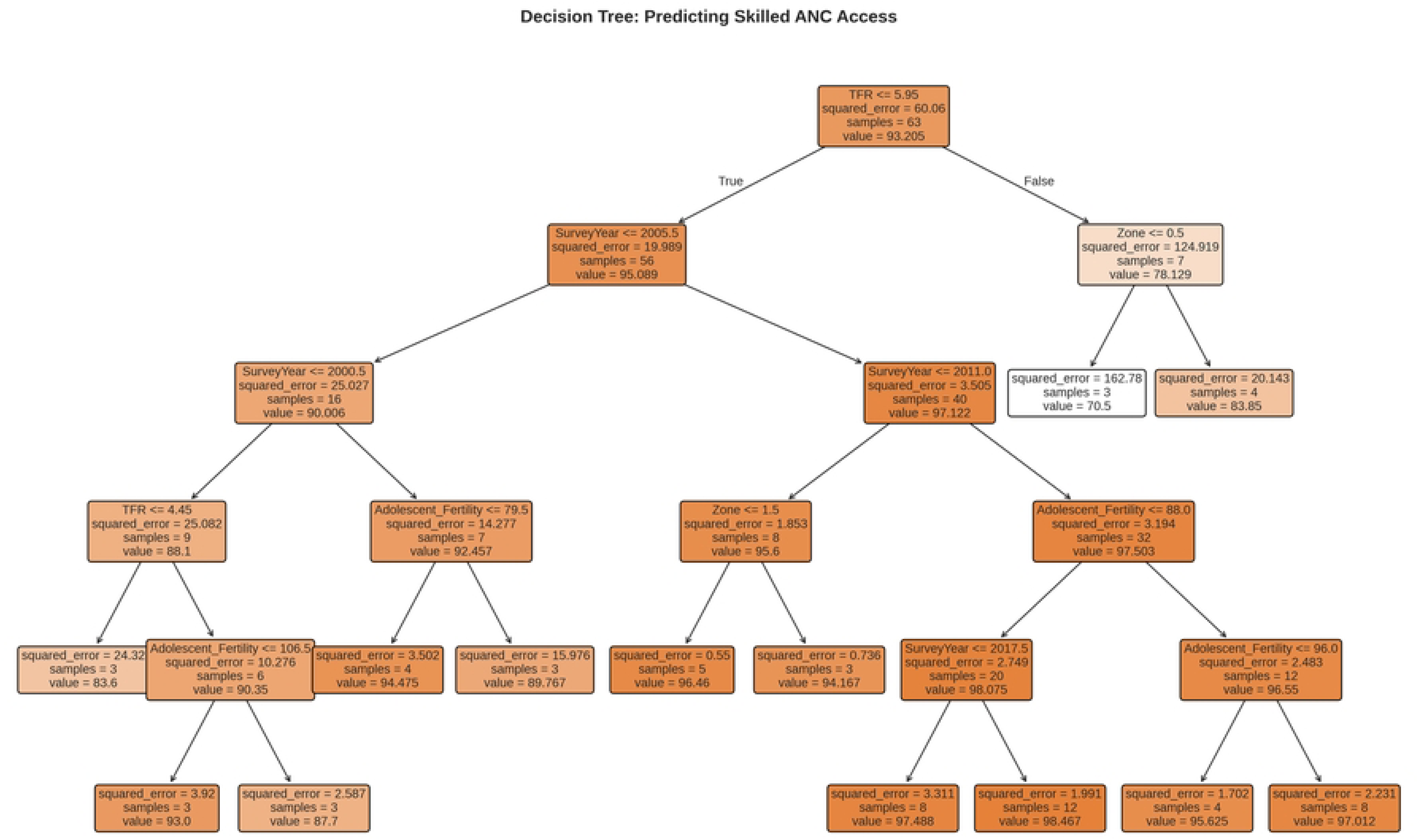
North–South ANC coverage gap by geographic belt, Ghana 1988–2022. Upper panel: mean ANC coverage by belt (Northern = red; Middle = gold; Southern = green; national average = blue dashed). Lower panel: South−North coverage gap in percentage points. Shaded area indicates North–South divergence zone.

**Fig 5.**
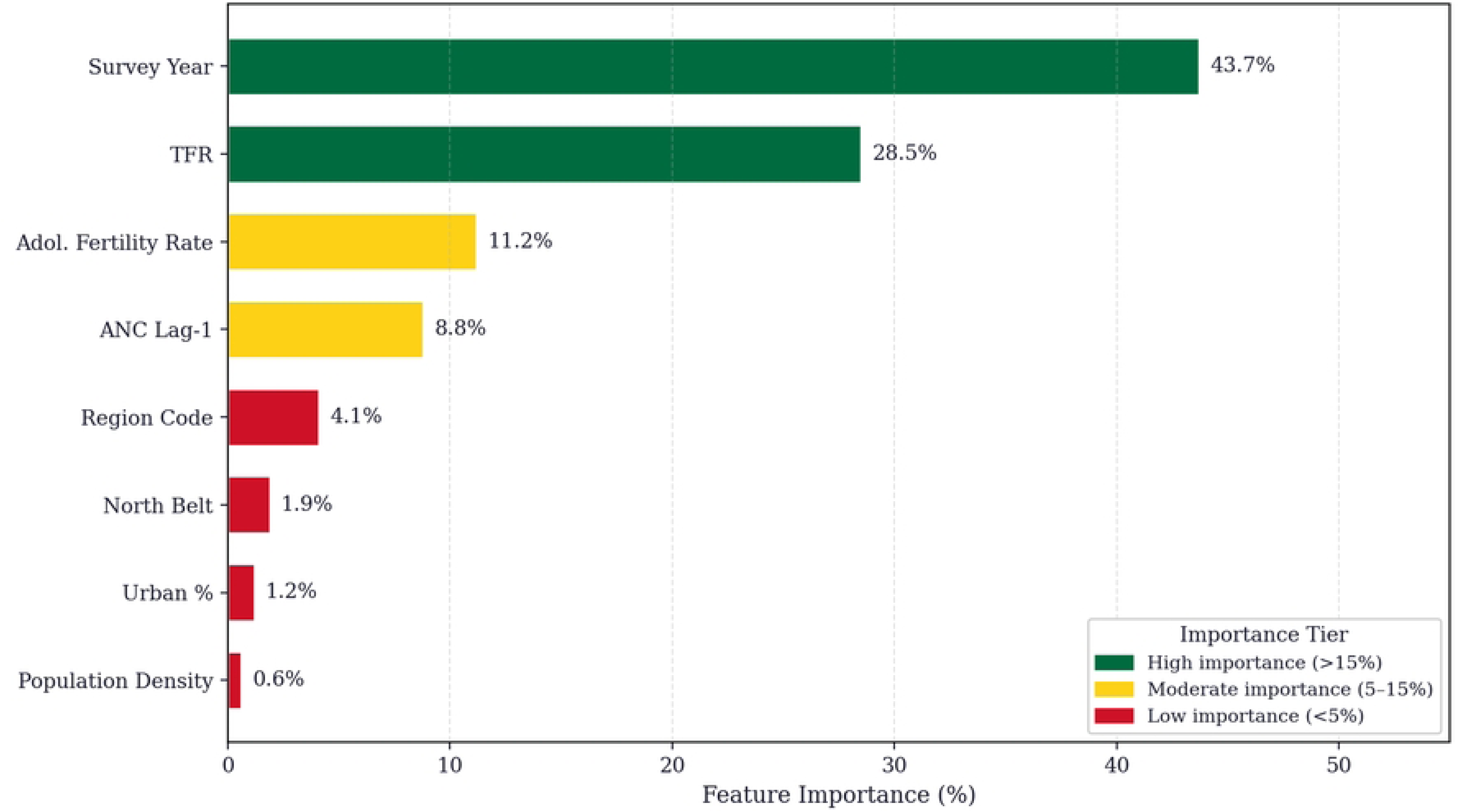
Inter-regional Gini coefficient for ANC coverage (green, solid) and total fertility rate (red, dashed), Ghana 1988–2022. Values annotated above each data point for ANC Gini. Declining ANC Gini reflects progressive convergence toward universal coverage.

### 3.3 Phase III: Machine Learning, Feature Attribution, and Risk Stratification

Out-of-sample generalization testing (RF Test R2 = 0.381; DT Test R2 = -0.296) demonstrates the Random Forest (RF) regressor’s substantial superiority over the Decision Tree (DT; Fig 6). The negative DT Test R2 shows that the single tree model predicted out-of-sample results worse than a mean-only baseline, indicative of overfitting given a small complete case training data set. Consequently, the DT is better described as a benchmark model. Predictive validity of the RF model is further demonstrated by five-fold cross-validation (CV R2 = 0.203; RMSE = 5.22%; MAE = 3.64%). Feature importance and RF analysis indicates that survey year is the leading RF predictor (43.7%) followed by TFR (38.8%), geographic zone (11.0%) and adolescent fertility rate (6.5%) (Fig 7). The DT is attributed as giving greater importance to TFR by itself (63.1%), which demonstrates the known divergence of variance attribution of ensemble and single tree models [10].

**Fig 6.**
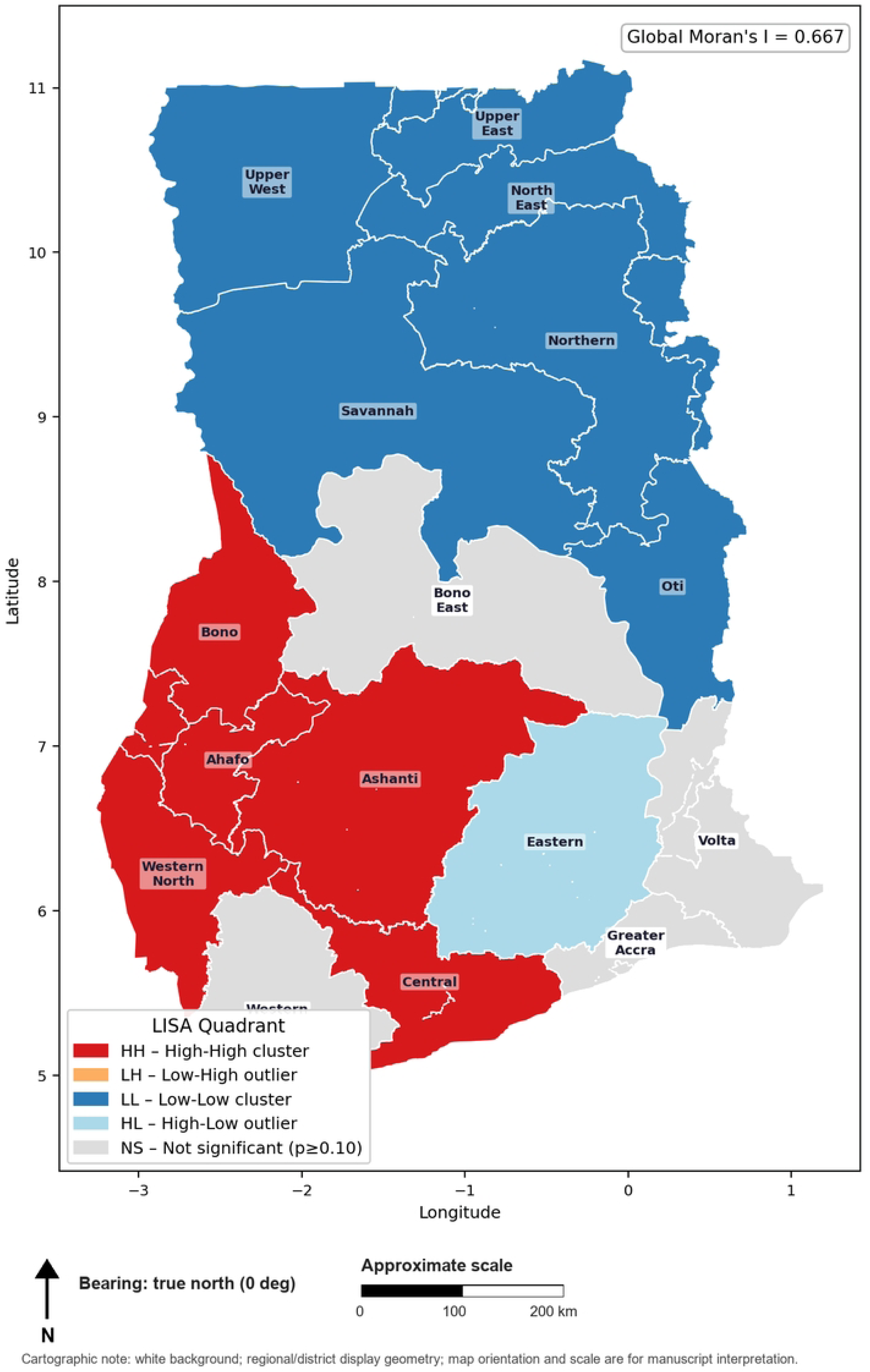
Decision Tree regressor structure for predicting skilled ANC coverage (single tree, max depth as fitted). Node splits show the predictor and threshold value; leaf nodes show predicted ANC coverage. Presented as a benchmark comparator to the Random Forest ensemble (Table 3).

**Fig 7.**
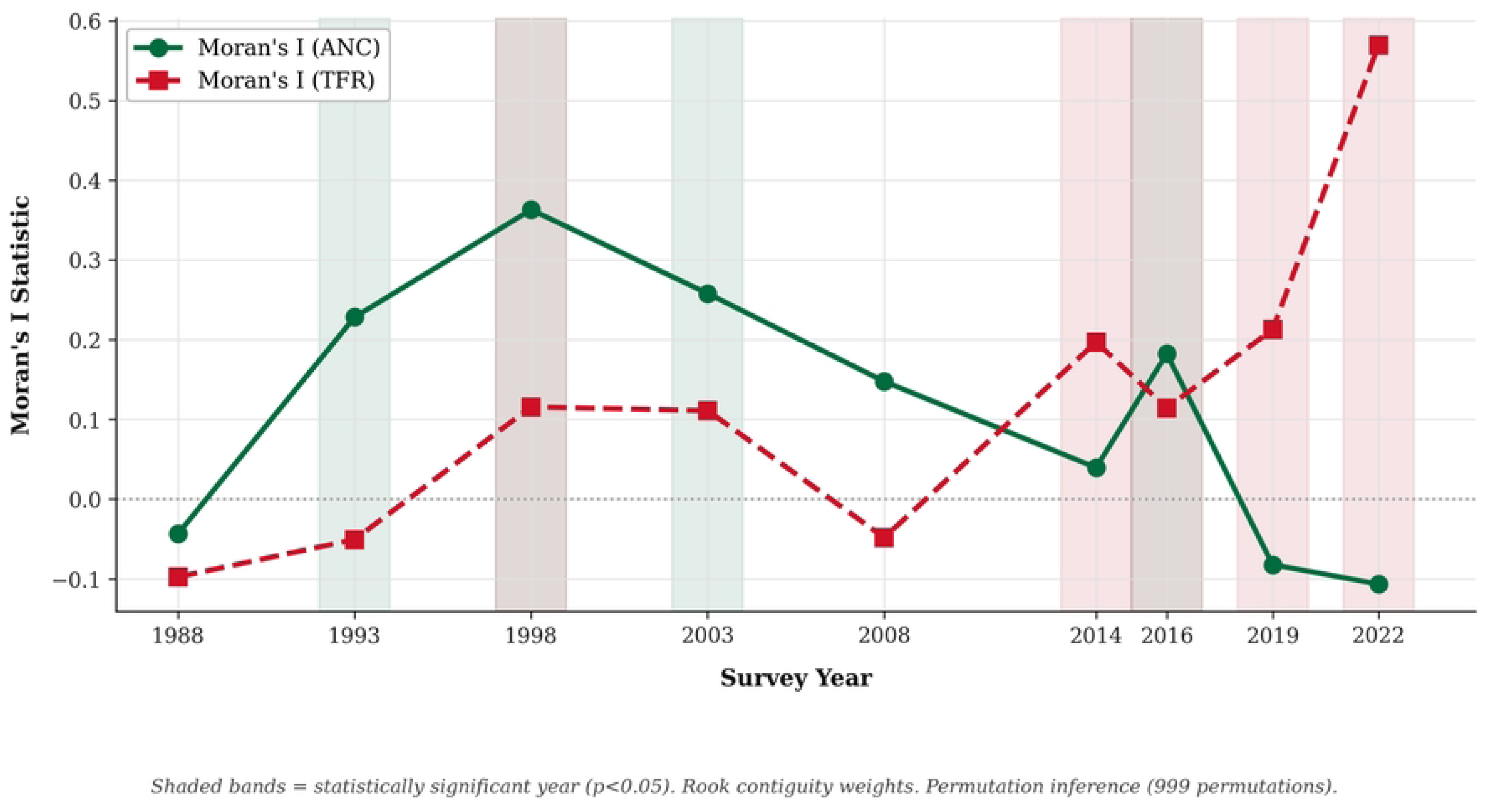
Feature importance for skilled ANC coverage prediction: Random Forest (200 trees, bars coloured by importance tier: green = high >15%, gold = moderate 5–15%, red = low <5%) and Decision Tree. Importance expressed as percentage of total variance explained.

**Table 3.**
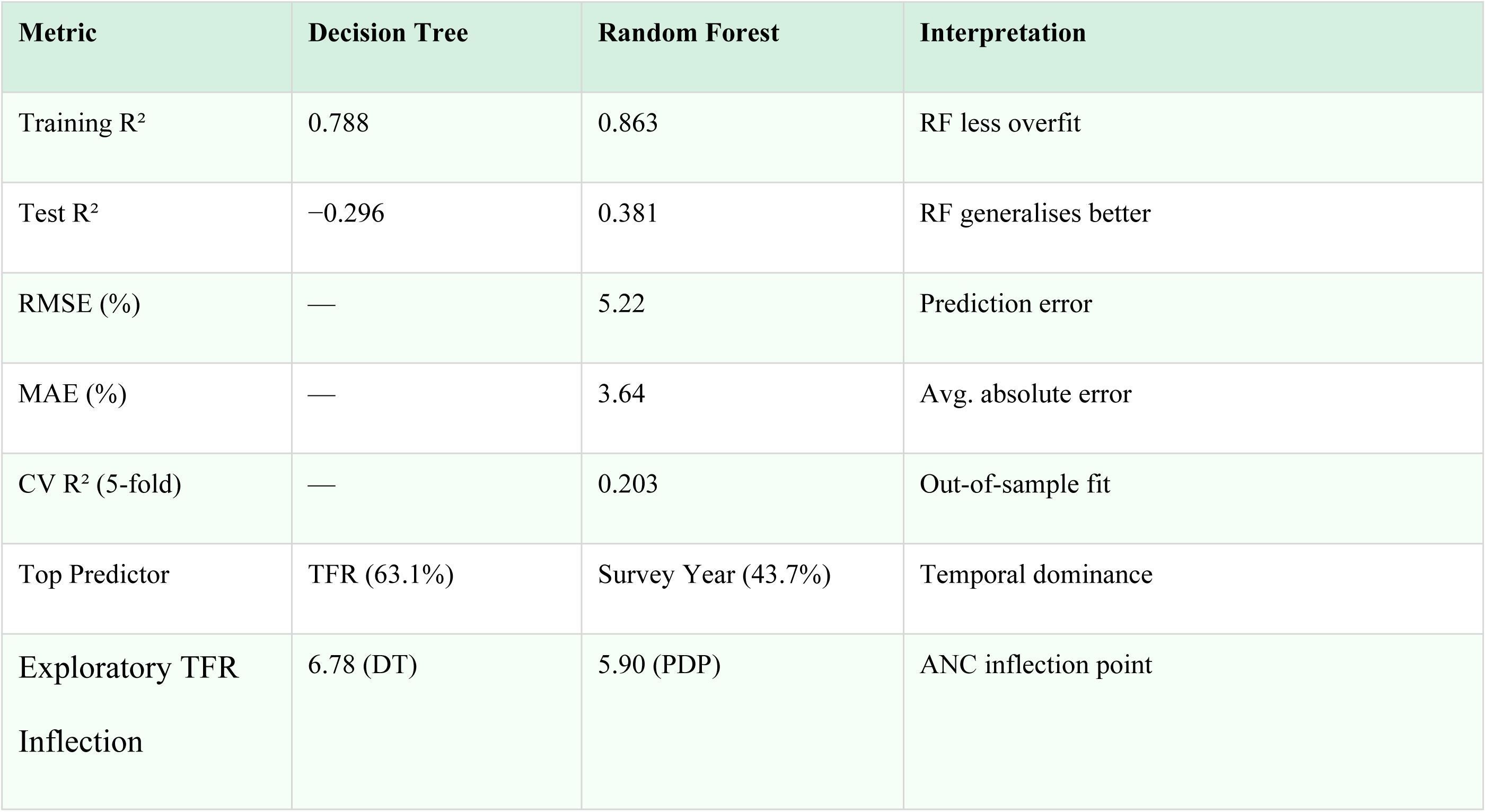
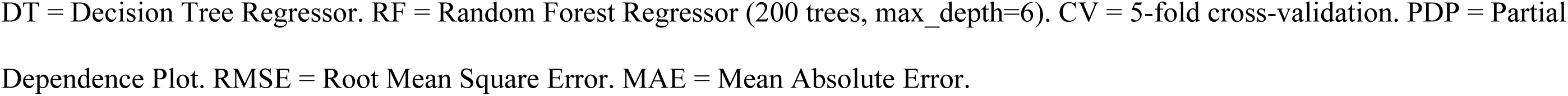
Machine Learning Model Performance Summary.

| Metric | Decision Tree | Random Forest | Interpretation |
| --- | --- | --- | --- |
| Training R<sup>2</sup> | 0.788 | 0.863 | RF less overfit |
| Test R<sup>2</sup> | −0.296 | 0.381 | RF generalises better |
| RMSE (%) | — | 5.22 | Prediction error |
| MAE (%) | — | 3.64 | Avg. absolute error |
| CV R<sup>2</sup> (5-fold) | — | 0.203 | Out-of-sample fit |
| Top Predictor | TFR (63.1%) | Survey Year (43.7%) | Temporal dominance |
| Exploratory TFR Inflection | 6.78 (DT) | 5.90 (PDP) | ANC inflection point |

DT = Decision Tree Regressor. RF = Random Forest Regressor (200 trees, max\_depth=6). CV = 5-fold cross-validation. PDP = Partial Dependence Plot. RMSE = Root Mean Square Error. MAE = Mean Absolute Error.

An exploratory TFR inflection is posited in the range of 5.90 based on partial dependence analysis (Fig 8). TFR levels above this inflection are associated with a predicted decline in skilled ANC coverage in the historical panel decaying from approximately 95-100% to 85-90%. This is hypothesized to indicate a limit rather than a policy restriction or a causal relationship. Table 4 presents the Care Efficiency Index by region and is pooled across all survey years.

**Fig 8.**
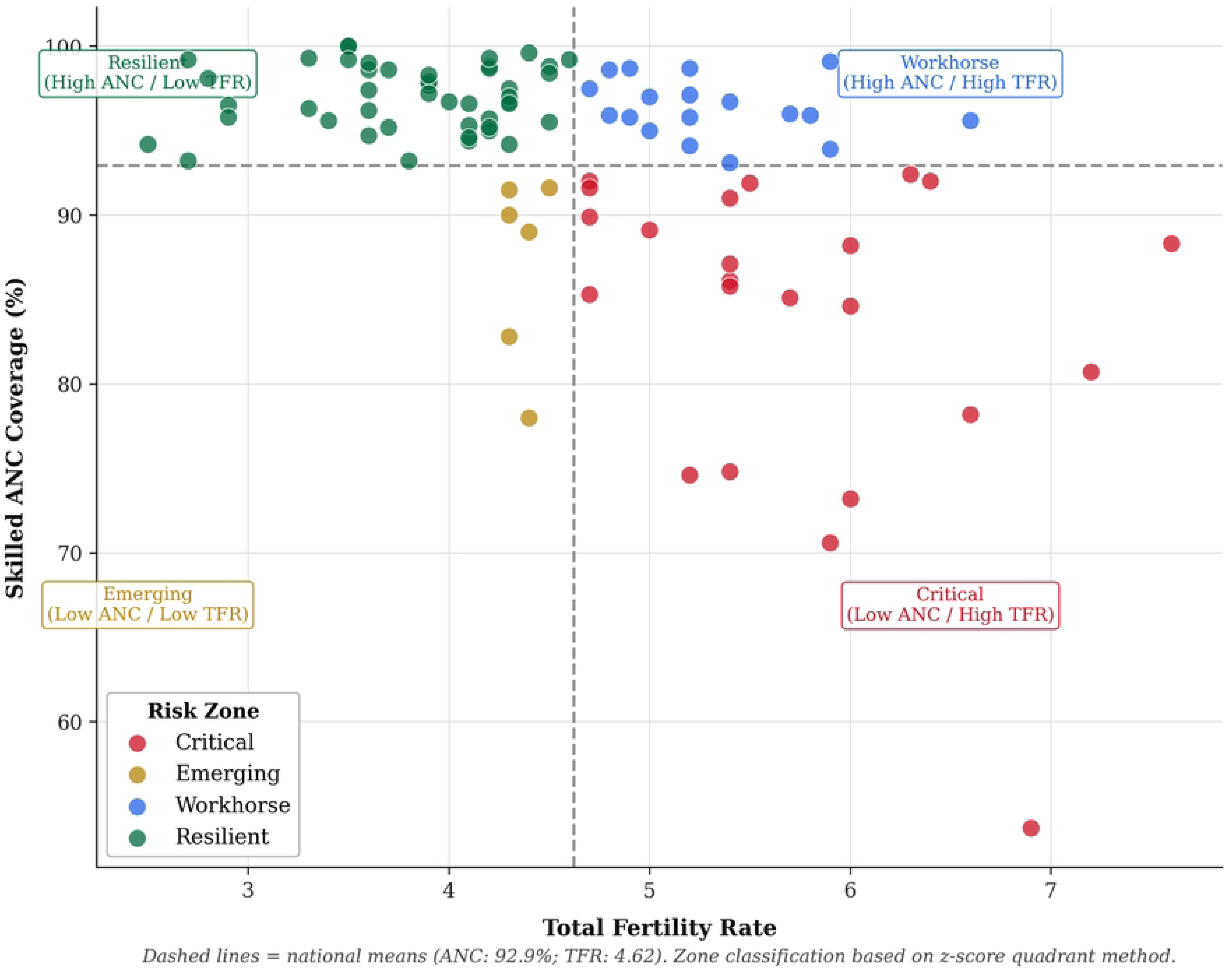
Partial dependence of predicted skilled ANC coverage on TFR (Random Forest regressor, 200 trees). Red dashed vertical line = exploratory RF partial-dependence inflection near TFR=5.90. Above this point, predicted ANC coverage declines in a non-linear pattern in the historical panel.

**Table 4.** Care Efficiency Index (CEI) by Region — Pooled Average 1988–2022.

| Rank | Region | Skilled<br>ANC (%) | TFR | CEI | Tier |
| --- | --- | --- | --- | --- | --- |
| 1 | Greater Accra | 95.6 | 3.10 | 31.9 | High |
| 2 | Bono | 98.6 | 3.70 | 26.6 | High |
| 3 | Western North | 98.6 | 3.80 | 25.9 | High |
| 4 | Eastern | 93.9 | 4.22 | 23.2 | High |
| 5 | Ashanti | 96.4 | 4.38 | 23.0 | High |
| 6 | Western | 94.2 | 4.48 | 21.8 | High |
| 7 | Bono East | 99.4 | 4.70 | 21.1 | High |
| 8 | Volta | 90.4 | 4.62 | 20.4 | Middle |
| 9 | Upper East | 92.6 | 4.62 | 20.3 | Middle |
| 10 | Ahafo | 95.6 | 4.91 | 20.2 | Middle |
| 11 | Central | 92.8 | 4.81 | 20.1 | Middle |
| 12 | Oti | 97.1 | 5.20 | 18.7 | Middle |
| 13 | Upper West | 90.6 | 4.99 | 18.5 | Middle |
| 14 | Savannah | 95.9 | 5.80 | 16.5 | Middle |
| 15 | Northern | 85.1 | 5.82 | 14.8 | Priority |
| 16 | North East | 95.6 | 6.60 | 14.5 | Priority |

CEI = Care Efficiency Index = Skilled ANC Coverage (%) ÷ Total Fertility Rate. Higher CEI = greater care coverage relative to fertility burden.

Bivariate risk stratification categorized 23 region-year observations as Critical (Low ANC / High TFR; 24.5%), 46 as Resilient (48.9%), 19 as Workhorse (20.2%), and 6 as Emerging (6.4%) (see Appendix Table 5; Fig 9). Critical observation clusters mainly occupied the early survey waves of the Northern Belt. As of 2022, most regions had moved beyond the Low ANC/High TFR classification, while the Northern Belt retained high fertility Workhorse patterns. Trends in the adolescent fertility rate by belt are illustrated in Fig 10.

**Fig 9.**
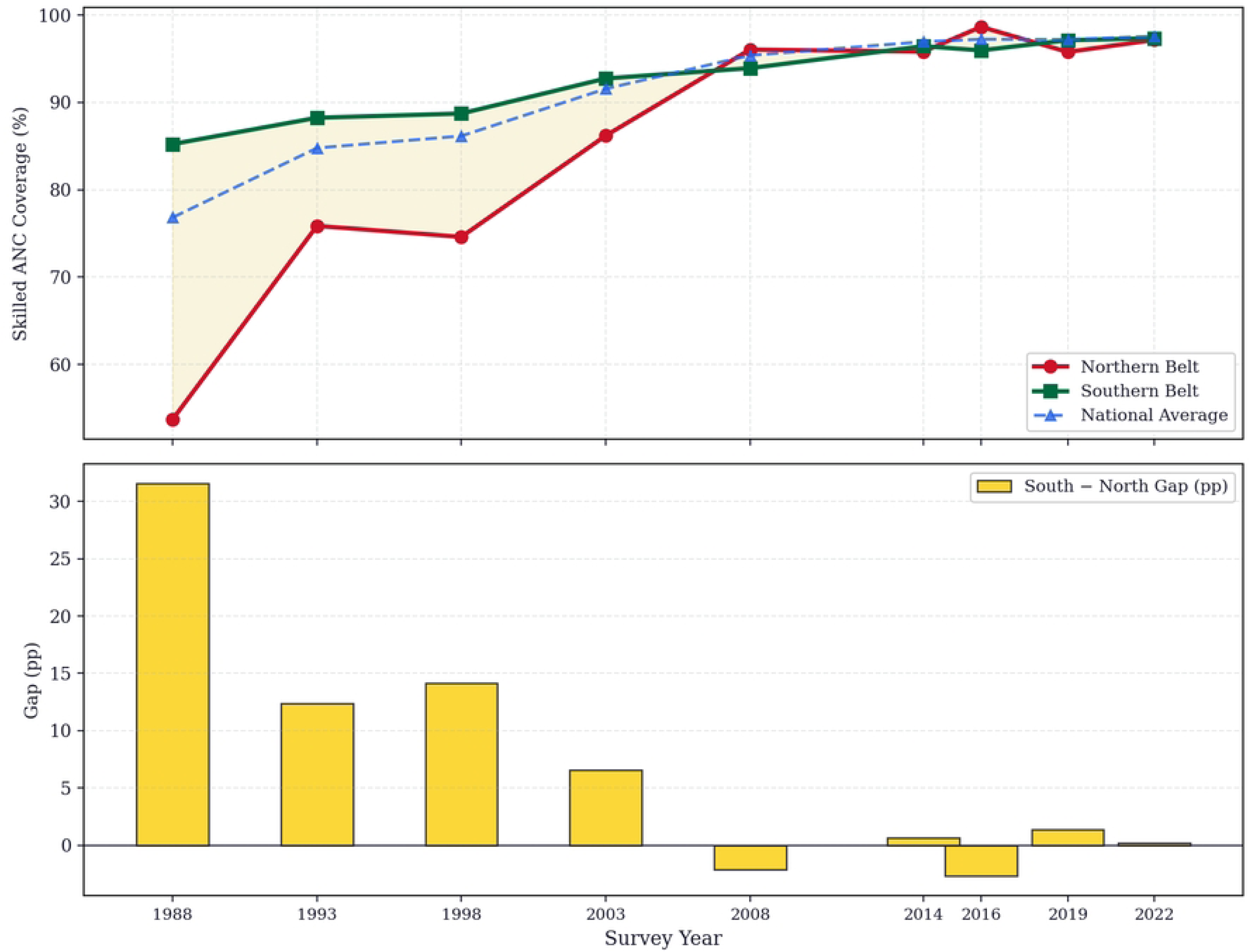
Bivariate ANC–TFR risk stratification dashboard, Ghana 1988–2022. Each data point represents one region-year observation. Dashed crosshairs = grand means. Zone colours: red = Critical (Low ANC/High TFR), gold = Emerging, blue = Workhorse, green = Resilient. Bar charts show zone counts and trajectory by survey year.

**Fig 10.**
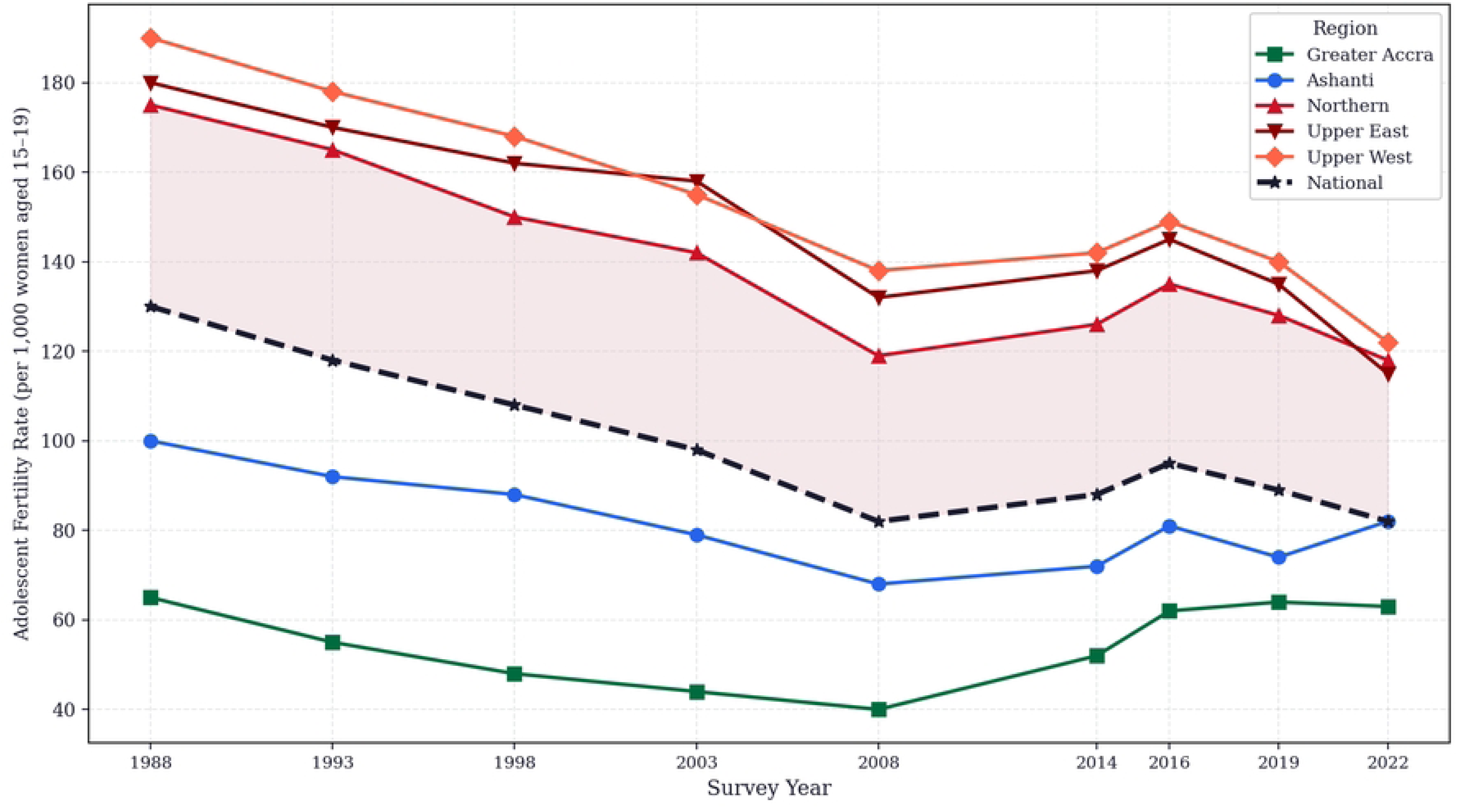
Adolescent fertility rate (births per 1,000 women aged 15–19) by region, Ghana 1988–2022. Northern Belt regions (red tones) maintain substantially higher adolescent fertility throughout the study period. National average shown as black dashed line.

**Table 5.** Bivariate Risk Zone Stratification Summary, Ghana 1988–2022.

| Risk Zone | N Obs. | Mean<br>ANC (%) | Mean TFR | Mean Adol.<br>Fert. | Mean CEI |
| --- | --- | --- | --- | --- | --- |
| Critical<br>(Low ANC /<br>High TFR) | 23 | 83.7 | 5.74 | 112.5 | 14.9 |
| Emerging<br>(Low ANC /<br>Low TFR) | 6 | 87.2 | 4.37 | 82.0 | 20.0 |
| Workhorse<br>(High ANC /<br>High TFR) | 19 | 96.5 | 5.28 | 93.5 | 18.4 |
| Resilient<br>(High ANC /<br>Low TFR) | 46 | 97.0 | 3.81 | 67.8 | 26.1 |

Zone classification based on z-score quadrants (ANC z-score vs TFR z-score). N Obs = number of region-year observations. Adol. Fert. = adolescent fertility rate (15–19 years) per 1,000. CEI = Care Efficiency Index.

### 3.4 Phase IV: Spatial Autocorrelation and LISA Analysis

Global Moran’s I analysis (Table 6, Fig 11) showed significant positive spatial autocorrelation in ANC coverage during the early-to-middle survey period (1993: I=0.228, p=0.020; 1998: I=0.363, p=0.008; 2003: I=0.258, p=0.016). This indicates geographic clustering of both high and low ANC coverage. By 2022, ANC clustering was non-significant (I=-0.042, p=0.395), whereas TFR clustering was strong and significant (I=0.606, p=0.001). This divergence suggests decoupling between supply-side improvements in health service coverage and the demographic and cultural determinants of fertility, which respond over longer time horizons.

**Fig 11.**
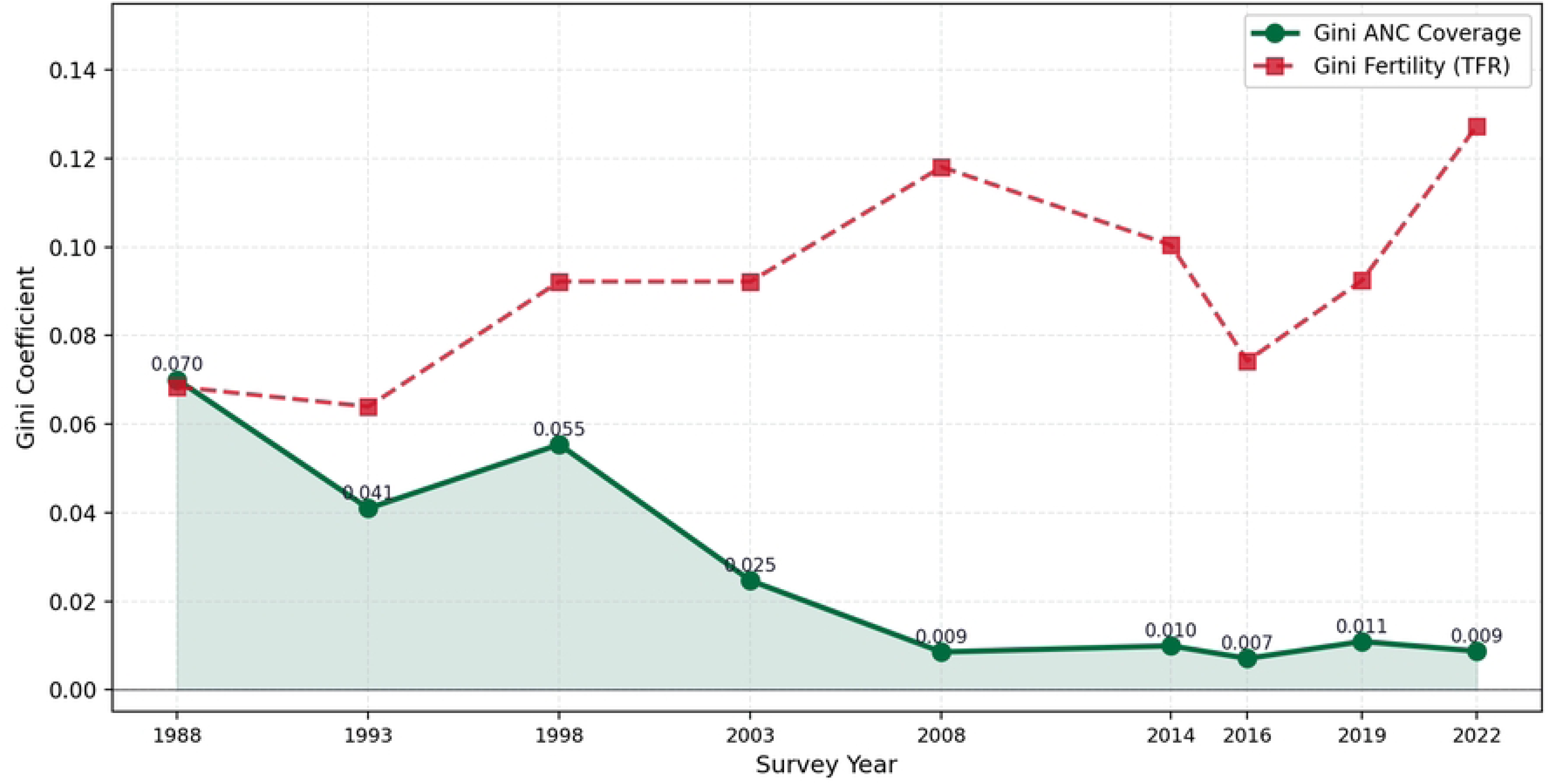
Temporal evolution of Moran’s I spatial autocorrelation for ANC coverage (green) and TFR (red), Ghana 1988–2022. Filled/bolded markers indicate p<0.05 (999 permutations, KNN k=4). Divergence of ANC and TFR autocorrelation by 2022 is clearly visible.

**Table 6.** Global Moran’s I Spatial Autocorrelation Statistics, Ghana 1988–2022.

| Year | Moran's I<br>(ANC) | p-value<br>(ANC) | Moran's I<br>(TFR) | p-value<br>(TFR) | n Regions | Interpretation |
| --- | --- | --- | --- | --- | --- | --- |
| 1988 | −0.044 | 0.158 | −0.098 | 0.299 | 8 | Random |
| 1993 | 0.228 | 0.020 | −0.051 | 0.268 | 10 | ANC<br>Clustered* |
| 1998 | 0.363 | 0.008 | 0.116 | 0.049 | 10 | Clustered* |
| 2003 | 0.258 | 0.016 | 0.111 | 0.051 | 10 | ANC<br>Clustered* |
| 2008 | 0.148 | 0.059 | −0.048 | 0.278 | 10 | Random |
| 2014 | 0.039 | 0.134 | 0.197 | 0.022 | 10 | TFR<br>Clustered* |
| 2016 | 0.182 | 0.024 | 0.114 | 0.026 | 10 | Clustered* |
| 2019 | −0.082 | 0.354 | 0.213 | 0.016 | 10 | TFR<br>Clustered* |
| 2022 | −0.042 | 0.395 | 0.606 | 0.001 | 16 | TFR<br>Clustered*** |

\* $p < 0.05$ \*\* $p < 0.01$ . KNN spatial weights ( $k=4$ ). Inference based on 999 permutations; the smallest attainable permutation p-value is 0.001.

LISA analysis (Figs 12, 13) indicated persistent Northern Belt spatial structure in earlier waves and a progressive attenuation of ANC clustering by 2022. By contrast, TFR clustering intensified by 2022, indicating that fertility-related spatial inequity remained patterned even as coverage converged. These findings are consistent with Ghana-specific evidence linking Northern Belt ANC underutilisation and fertility patterns to transportation constraints, cultural norms, female educational opportunity, and health workforce distribution [4,5].

**Fig 12.**
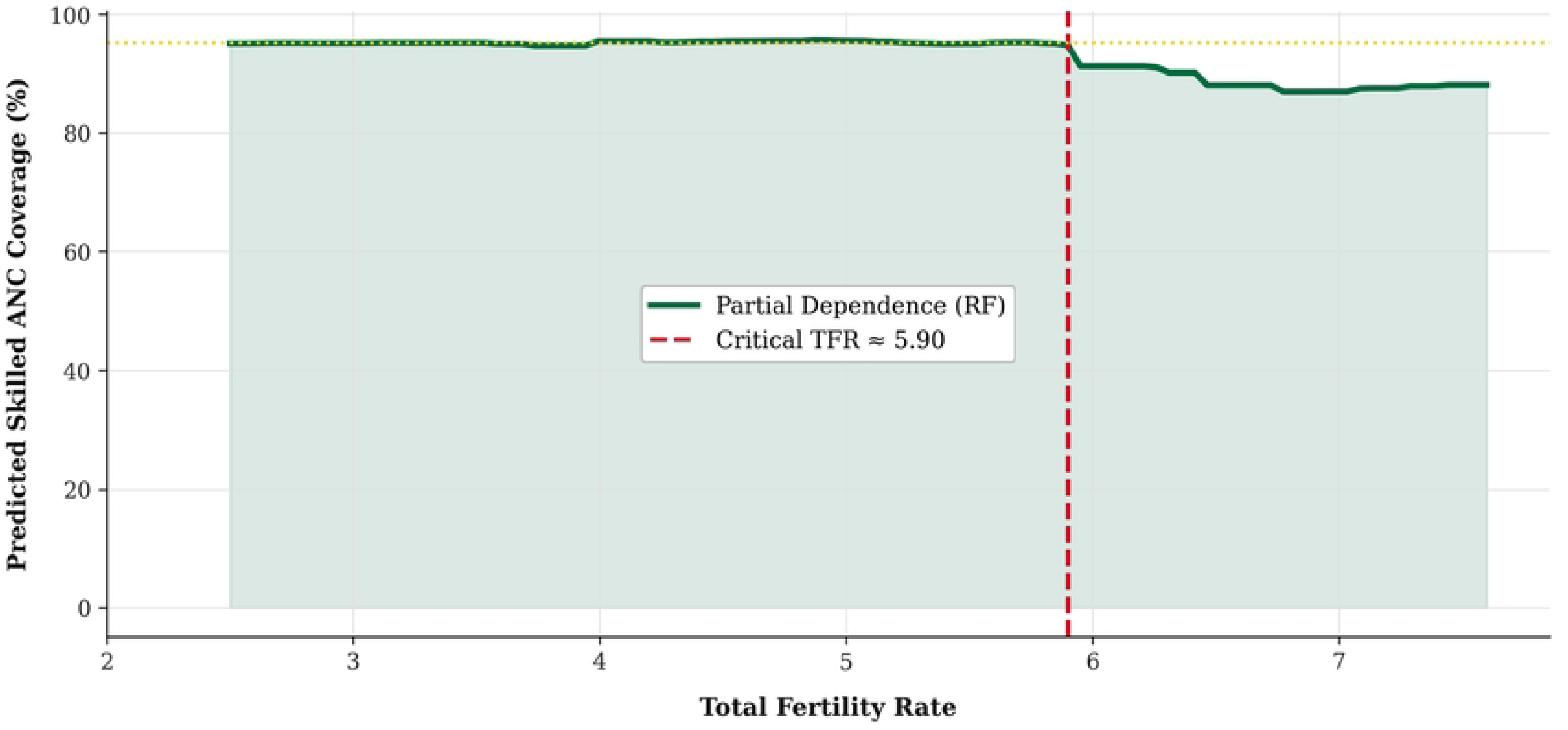
LISA cluster map of skilled ANC coverage by region, pooled 1988–2022 (Local Moran’s I, p<0.10 significance threshold). HH (red) = High-High cluster; LL (blue) = Low-Low cluster; HL (orange) = High-Low outlier; NS (grey) = not significant. Global Moran’s I annotated. Source: Ghana DHS 1988–2022 [3,13]. Random Forest (200 trees, max depth 6). Critical TFR threshold = 5.90 (maximum gradient change in predicted ANC). All other features held at sample mean.

**Figure 13:**
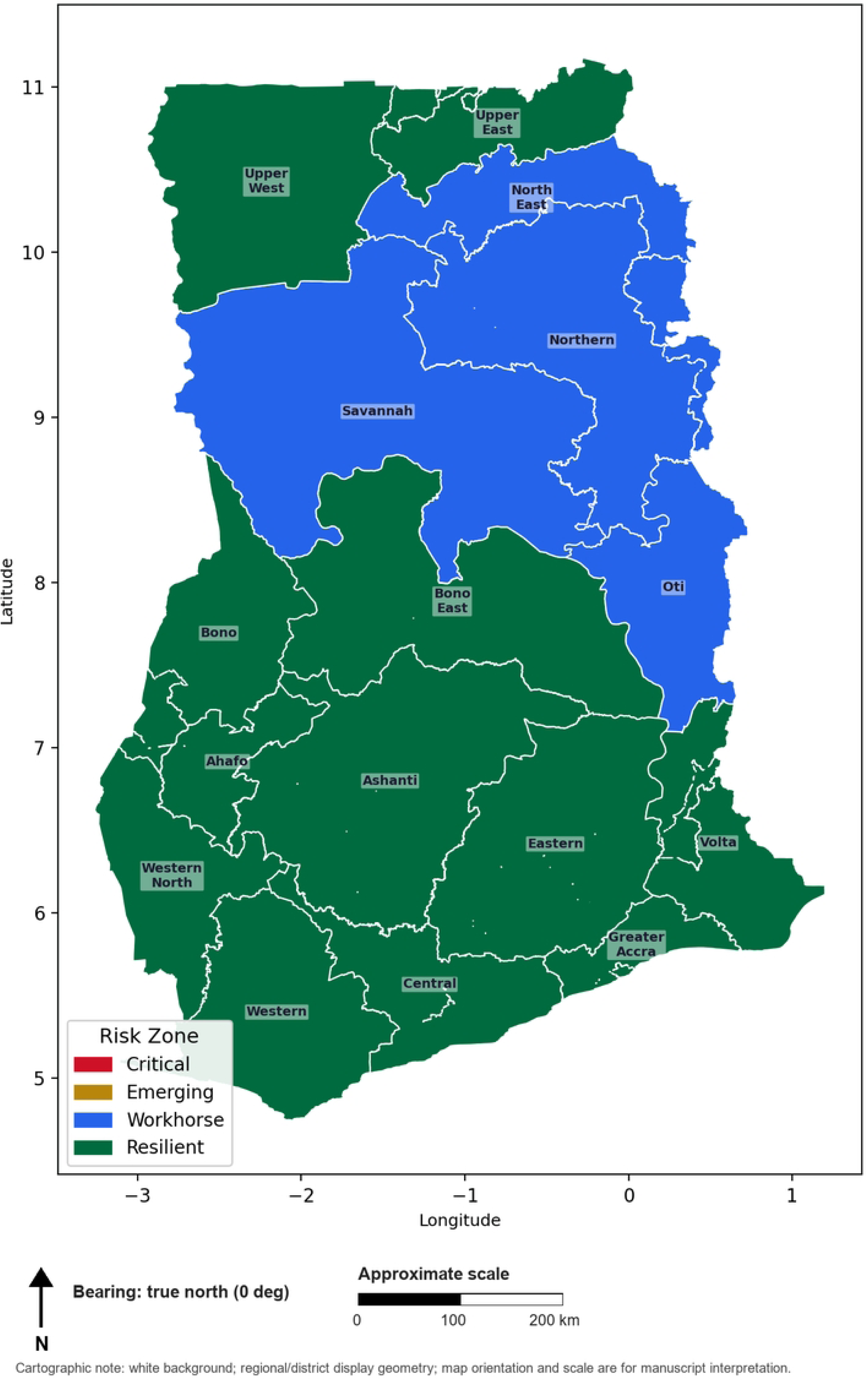
Bivariate Risk Zone Choropleth by Region, Ghana, 2022. Critical = Low ANC and High TFR (red). Emerging = Low ANC and Low TFR (gold). Workhorse = High ANC and High TFR (blue). Resilient = High ANC and Low TFR (green). Source: Ghana DHS 2022 [3,13].

### 3.5 Supplementary Maps: Regional ANC, TFR, CEI, Risk Zone, and Temporal Choropleth

Figure 14 shows almost total coverage of Skilled ANC in all regions in 2022, with Greater Accra (95.8%), Eastern (99.2%) and Ashanti (100%) achieving high levels. The coverage in the Savannah (95.9%) and Northern (96.7%) Regions, while significantly improved, now places them below the regional mean. The TFR for 2022 (Fig. 15) continues to illustrate the North-South divide. In the North, the Savannah (TFR 5.8), Northern (TFR 5.4) and North East (TFR 6.6) Regions all have TFRs that are significantly higher than the regional mean. In the South, Greater Accra (TFR 2.9) and Eastern (TFR 3.5) have TFRs that are significantly lower. In the CEI choropleth (Fig. 16), Greater Accra stands out as the efficiency leader (CEI = 31.9), while North East is the laggard region (CEI = 14.5), reflecting a 2.2 times efficiency gap despite a convergence in coverage. The temporal choropleth (Fig.17) shows the coverage of Skilled ANC.

**Figure 14:**
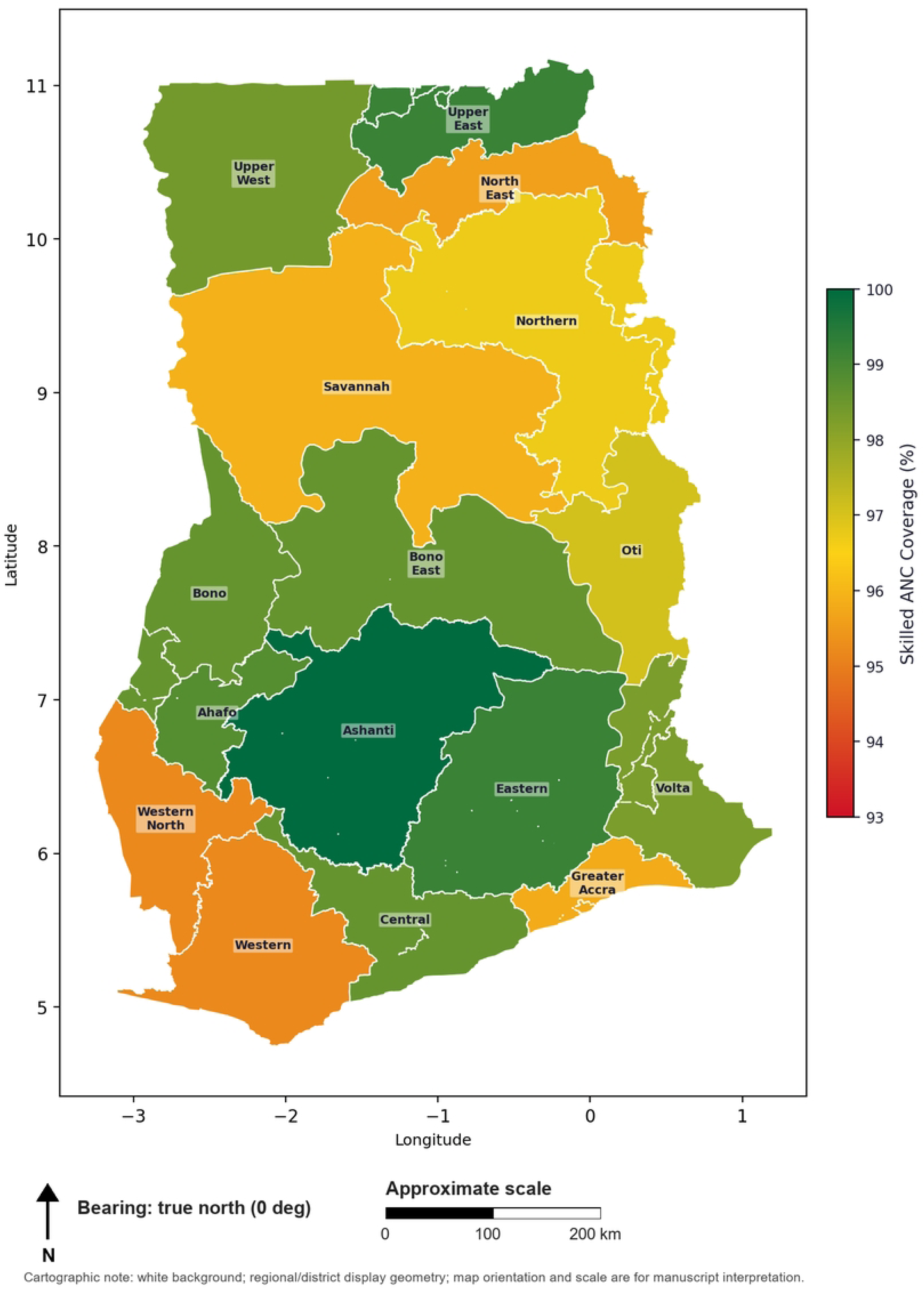
Skilled ANC Coverage (%) by Region, Ghana, 2022, in Choropleth Format. Red = low coverage, green = high coverage (shown in color bar). Regions are the 16 administrative areas of Ghana. Source: Ghana DHS 2022 [3,13].

**Figure 15:**
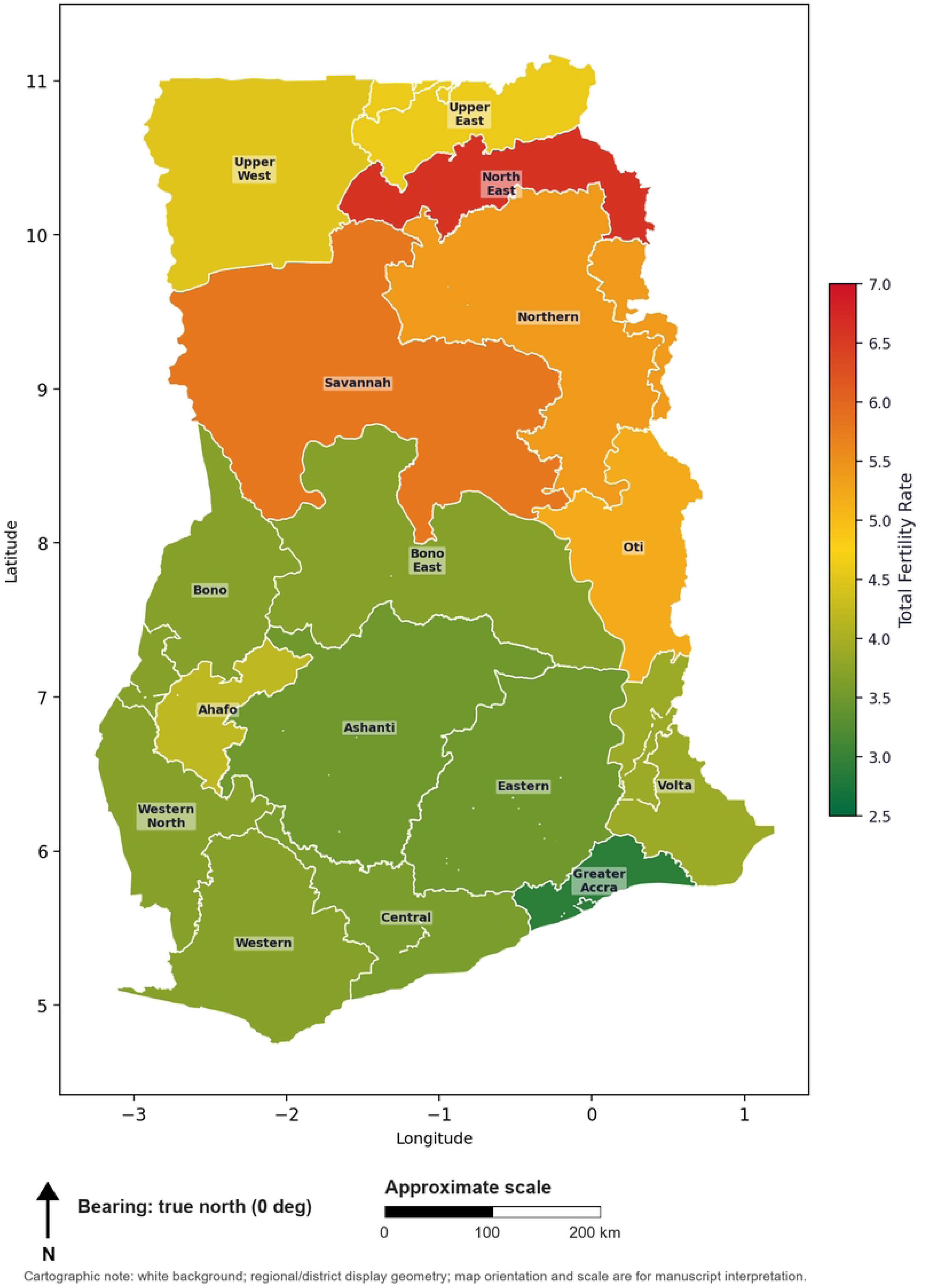
Total Fertility Rate (TFR) in Region-Based Choropleth Mapping, Ghana 2022. Green = lower TFR, red = higher TFR (shown in color bar). Persistent North-South gradient. Source: Ghana DHS 2022 [3,13].

**Figure 16:**
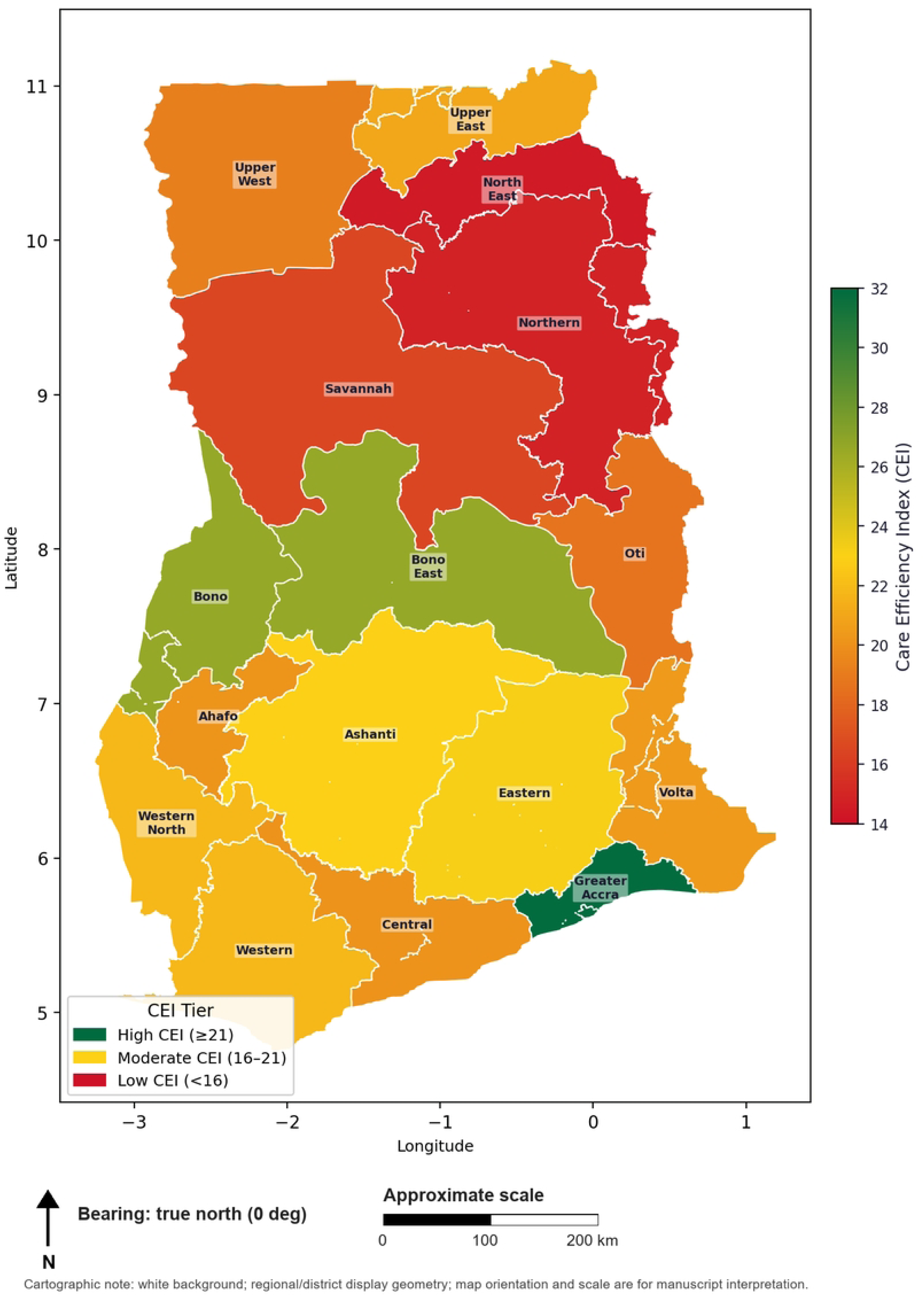
Care Efficiency Index (CEI = Skilled ANC% / TFR) by Region (pooled 1988-2022). Color bar: green = high efficiency, red = low efficiency. Tier Legend: High ≥21, Moderate 16-21, Low <16.

**Figure 17:**
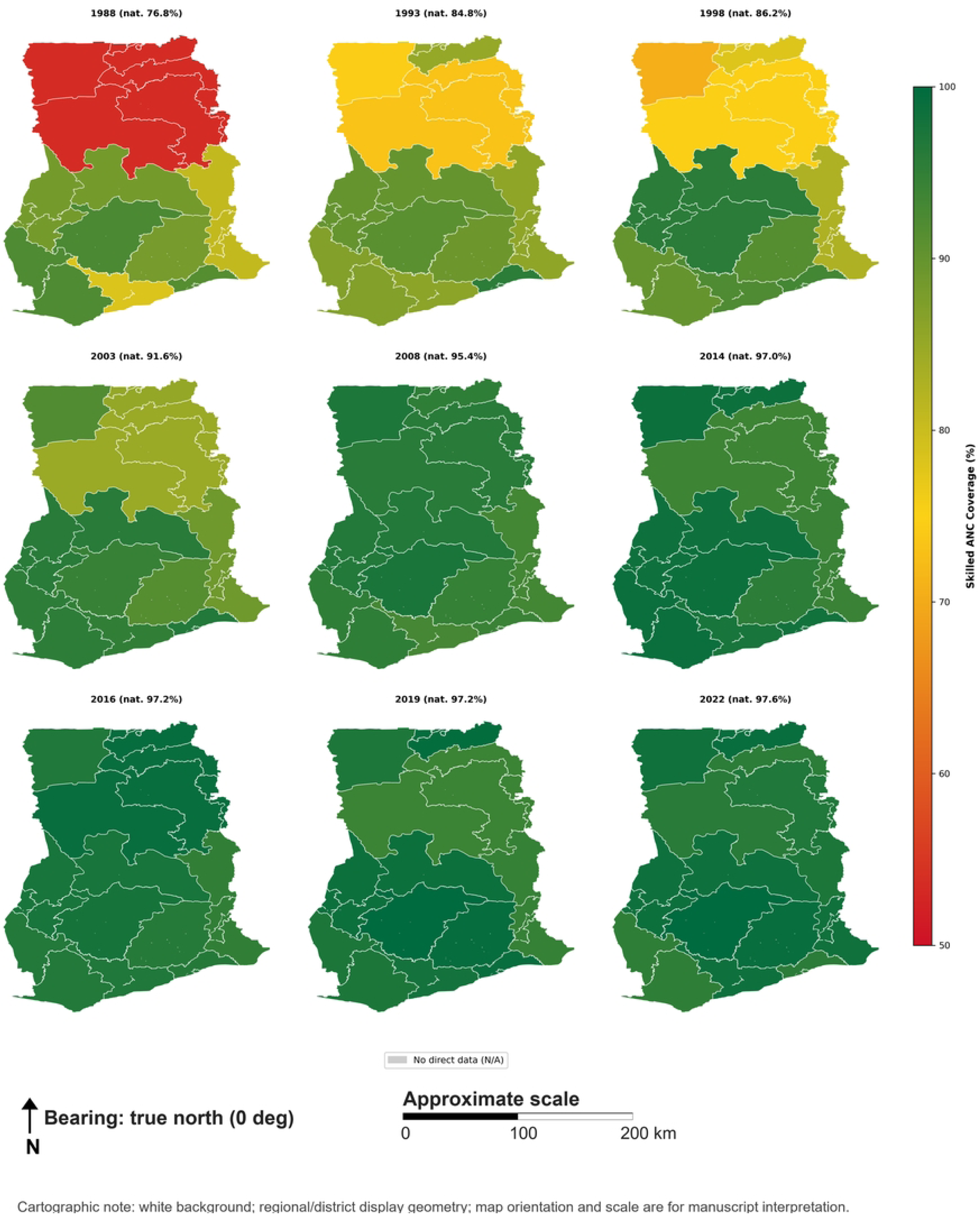
Temporal Choropleth Maps of Skilled ANC Coverage (%) by Region, Ghana, 1988-2022 (Nine Panels). Color scale is red (low) to green (high) in all panels and is therefore directly comparable. Grey = data not available.

### 4.1 Principal Findings

This study examines trends in Ghanaian coverage of skilled Antenatal Care (ANC) and fertility-related inequalities in and across nine Demographic and Health Survey (DHS) waves over the past 34 years [3, 13]. Four key findings emerge. First, ANC convergence was large (Gini reduced by 87.9%) and the Northern Belt’s fertility inequities remained hidden by large ANC coverage gains. Second, Total Fertility Rate (TFR) spatial clustering intensified in 2022 (Moran’s I=0.606; Table 6) while ANC clustering diminished indicating a significant cleaving of the two indicators. Third, an exploratory ANC inflection near 5.90 indicated a sharp decline in ANC and needs to be validated prior to influencing policy. Finally, the Care Efficiency Index demonstrated a 2.2 fold performance gap between Greater Accra and North East, which would be obscured by coverage rates.

### 4.2 ANC Convergence and the Persistence of Fertility Gradients

The near-elimination of inter-regional ANC inequality (Gini 0.070 to 0.008; Table 2, Fig 5) coincided with Ghana’s National Health Insurance Scheme introduction in 2003, the abolition of user fees for facility-based delivery services in 2003, and subsequent community health worker expansion [14,15,29]. Causal attribution cannot be made from this ecological design [30]. However, the timing and direction of change are consistent with policy-driven service expansion. The persistence of high fertility in Northern Belt regions (North East TFR=6.6; Savannah TFR=5.8; Northern TFR=5.4) relative to Greater Accra (TFR=2.9) shows that fertility transition remains incomplete in the Northern Belt [3,28]. ANC coverage convergence alone therefore does not guarantee fertility-related equity.

This finding extends prior work on health insurance non-enrolment in Ghana [6] and aligns with evidence that demographic and health disadvantages in the Northern Belt are structurally reinforced across multiple domains [4,5,17].

### 4.3 Machine Learning and the Exploratory TFR Inflection

Prior to the implementation of a policy, the exploratory TFR inflection at 5.90 (Table 3, Fig 8) must be validated with a larger, region-stratified sample. The RF model demonstrated a moderate fit (CV R2=0.203). A train/test split, along with survey-year stratification, may have introduced temporal leakage. Many of the Northern Belt regions in earlier survey waves were assigned predicted values of ANC coverage that were lower, given the specified contextual variables, which aligned with the RF model. This finding is aligned with the sociological evidence concerning the existence of high parity and/or opposition partners, as well as transportation and economic barriers, which may limit the demand for skilled antenatal care in the context of super-high fertility [4,7]. The RF estimate of 5.90, in comparison to the Decision Tree value of 6.78, indicates that the former is the result of ensemble smoothing over multiple paths along the tree [10].

The RF model placed the greatest importance on the survey year, indicating that the primary macro-level factors that explained the ANC improvements over the 34 years of the study were the secular time-related factors. These were likely the gradual strengthening of the health system, construction of health-related infrastructure, and the gradual expansion of the NHIS [15,29]. The RF model showed that TFR was 38.8% important, which indicates that the demographic context was most relevant to ANC coverage, especially when the survey year was taken into account..

### 4.4 Care Efficiency Index as a Policy Tool

To benchmark regional health system performance in a fertility-adjusted manner, the Care Efficiency Index (CEI = ANC% / TFR) can be used. It shows the performance of the regional health system in terms of the existing RMNCH coverage indicators and highlights the situations where high contact coverage and high fertility pressure exist. Greater Accra achieved the highest CEI (31.9; Table 4), while North East recorded the lowest (14.5). With assessment based on ANC coverage alone, this 2.2-fold gap would be missed..

### 4.5 Risk-Zone Stratification as a Spatial Targeting Tool

Bivariate risk stratification adds spatial and temporal specificity to the trends above. Of 94 region-year observations, 23 (24.5%) met Critical criteria (Table 5, Fig 9) (low ANC coverage / high TFR), concentrated in the Northern Belt during earlier survey waves and consistent with the TFR inflection identified nationally (§4.3). By 2022, the plurality of observations (46/94, 48.9%) had transitioned to Resilient status, indicating that ANC gains reached the sub-regional level rather than remaining a national aggregate effect. However, Workhorse status (high ANC coverage coexisting with high TFR; 19/94, 20.2%) persisted in parts of the Northern Belt through 2022. Workhorse and Resilient regions both report adequate coverage, but a Workhorse classification implies a materially higher per-provider service burden — more antenatal contacts, deliveries, and commodity consumption relative to staffing — than a Resilient region achieving the same coverage under lower fertility pressure. This distinction is relevant to the resource-allocation recommendation in §5: differential planning based on risk-zone status should treat Workhorse regions as a staffing-intensity signal, not simply a coverage success. It was observed that pooled 1988–2022 zone counts describe population-level tendencies over the full study period and do not by themselves identify which regions require intervention now; a 2022-specific zone assignment per region would be needed before any district-level resource decision (§4.7).

Sensitivity analysis supported the use of risk zones as exploratory targeting strata rather than deterministic allocation categories. When risk-zone labels were recalculated against same-year rather than grand historical z-score baselines, 54.3% of observations retained the same classification, and Critical-zone overlap had a Jaccard index of 0.324. The 2022 same-year classification identified 4 Critical, 3 Workhorse, 2 Emerging, and 7 Resilient regions. Thus, the central finding of persistent subnational inequity was robust, but exact zone labels changed with the reference distribution.

### 4.6 Adolescent Fertility and Intergenerational Equity

Adolescent fertility rates (Fig 10) in the Northern Belt remained above the national average throughout the study period. Upper East and Upper West exceeded 160 births per 1,000 women aged 15-19 in 1988 and still exceeded 115 per 1,000 by 2022. Persistent adolescent fertility is both a proximate driver of high regional TFR and an indicator of unmet need for reproductive health services among young women [1,18]. Female educational attainment is inversely associated with maternal mortality and high fertility across global contexts [17], and remains lower in Ghana’s Northern Belt than in the south [3,28]. These patterns indicate that the equity challenge extends beyond the health sector.

### 4.7 Limitations

This analysis has several limitations. The ecological design precludes individual-level causal inference; regional associations between TFR and ANC coverage may be confounded by female educational attainment [17], road network density, health facility density [20,21], and other unmeasured structural factors. The analytical sample (n=94 region-year observations, with a smaller complete-case subset for machine learning) is small for predictive modelling, and the RF model achieved only moderate out-of-sample R2 (0.381). DHS subnational data also suppress within-region heterogeneity; women in urban enclaves within Northern Belt regions may differ substantially from the regional mean [5,19]. The Care Efficiency Index is an ecological composite and should be validated against individual-level data and routine service utilisation registers before formal adoption. In addition, the risk-stratification z-scores (§2.7) were computed against a grand mean and standard deviation pooling all 94 region-year observations; six regions (Bono, Bono East, North East, Oti, Savannah, Western North) contribute only a single 2022 observation each, while the remaining ten contribute 8–9 waves spanning 34 years of historical trend. This reference distribution is not fully exchangeable — the grand mean is disproportionately anchored by other regions’ historical low-ANC/high-TFR years that the six single-wave regions never independently experienced, which could shift their z-score classification in either direction. A wave-stratified sensitivity analysis (comparing each region only against other regions’ same-year values) was not performed in this iteration and is recommended before the risk-zone classification is used for resource-allocation decisions. Finally, all 16 modern administrative regions are now represented, but six regions created by the 2018 subdivision (Bono, Bono East, North East, Oti, Savannah, Western North) have only a single independently-measured wave (2022); their pre-2022 trajectory cannot be characterised, and spatial complete-case denominators vary by model.

Robustness analyses further support a cautious interpretation. The crude ANC-TFR association was negative (Pearson r=-0.534), and remained negative after survey-year adjustment and cluster-robust specifications. However, the coefficient attenuated under GEE and became non-significant after simultaneous region and year adjustment. The ANC-TFR findings should therefore be read as ecological spatial-temporal patterning and decoupling, not as evidence that fertility directly reduces skilled ANC use.

## 5. CONCLUSIONS

Ghana’s trajectory from wide regional ANC inequality in 1988 to near-universal coverage in 2022 is a major public health achievement consistent with NHIS expansion, fee-exemption policy, and community health worker scale-up. Yet persistent TFR spatial clustering in the Northern Belt (Moran’s I=0.606, p=0.001), the exploratory TFR inflection near 5.90, and the 2.2-fold Care Efficiency Index gap between Greater Accra and North East define an ongoing equity challenge that conventional ANC coverage statistics alone cannot reveal. Policy priorities include targeted fertility transition and female educational empowerment programmes in Northern Belt regions exceeding TFR 5.0, pilot validation of the CEI against DHIMS2 facility-level utilisation registers, spatial risk-zone mapping for differential resource allocation, and integration of spatial machine learning into Ghana Health Service annual district health data reviews. Future research should incorporate demand-side barriers and longitudinal facility-level data to separate supply from demand drivers of persistent ANC underutilisation in the Northern Belt.

## Data Availability

The main data sources for this research are Ghana’s Demographic and Health Surveys (1988-2022). The author does not hold proprietary rights to this data, nor can they redistribute it. The original data files for the surveys can be obtained at no cost from The DHS Program (https://dhsprogram.com) via registration and submission of a data access request. Level region indicators that are used here in aggregated form are also available for the DHS STATcompiler tool (https://www.statcompiler.com) and the DHS API (https://api.dhsprogram.com). The region-by-year analytic dataset and all the codes to perform the analysis and replicate the results have been published at Zenodo and can be viewed at this link (https://doi.org/10.5281/zenodo.21340351). The development repository for the analysis is available at GitHub (https://github.com/valentineghanem-bit/anc-fertility-ghana-inequities).

https://doi.org/10.5281/zenodo.21340351

## Declarations

### Ethics approval and consent to participate

This study used publicly available, de-identified, aggregate subnational data from the Ghana Demographic and Health Survey programme. No individual-level data were accessed. Institutional ethics review was not required per applicable guidelines for secondary analysis of publicly available anonymised data.

### Consent for publication

Not applicable.

## Data availability

The aggregate analytic dataset, code, bespoke HI-EI dashboard, and poster are available in the project repository: https://github.com/valentineghanem-bit/anc-fertility-ghana-inequities. Source DHS data are available from the DHS Program and STATcompiler subject to standard registration and data-use terms.

## Author contributions

Conceptualization: VGG. Data curation: VGG. Formal analysis: VGG. Investigation: VGG. Methodology: VGG. Software: VGG. Validation: VGG. Visualization: VGG. Writing – original draft: VGG. Writing – review & editing: VGG.

## Acknowledgements

The author acknowledges the DHS Program for making subnational health indicators publicly available, the Ghana Statistical Service for district boundary files, and the Ghana Health Service for the Annual Health Sector Performance Reports.

## Supporting Information

S1 File. Sensitivity analysis. Risk-zone classification sensitivity (grand-mean vs. same-year z-score baselines), ANC-TFR association robustness (HC3, cluster-robust, region and survey-wave fixed effects, GEE), and a descriptive bootstrap of the pooled regional Care Efficiency Index gap.

